# Genetic variants link lower segregation of brain networks to higher blood pressure and worse cognition within the general aging population

**DOI:** 10.1101/2021.08.12.21261975

**Authors:** Julia Neitzel, Rainer Malik, Ryan Muetzel, Maria J Knol, Hazel Zonneveld, Marios K Georgakis, Nicolai Franzmeier, Anna Rubinski, Martin Dichgans, M Arfan Ikram, Meike W Vernooij, Michael Ewers

## Abstract

The functional architecture of the brain is composed of distinct networks, where higher system segregation, i.e. greater differentiation of such functional networks, is associated with better cognitive performance. Aging and many neurological diseases have been associated with reduced system segregation and thus cognitive impairment. The genetic basis and risk factors of system segregation are largely unknown. Here, we present the first genome-wide association study of fMRI-assessed system segregation in 16,635 UK Biobank participants, identifying nine independent genomic loci. The 66 implicated genes were significantly downregulated in brain tissue and upregulated in vascular tissue. Of major vascular risk factors (Life’s Simple 7), blood pressure showed a robust genetic correlation with system segregation. Observational and Mendelian randomization analyses confirmed a unfavourable effect of higher blood pressure on system segregation and of lower system segregation on cognition. Replication analyses in 2,414 Rotterdam Study participants supported these conclusions.

## Introduction

The functional architecture of the brain is characterized by large-scale networks arising from neurovascular responses that are temporally coordinated between different brain regions^1^. These networks can be reproducibly detected during resting-state functional magnetic resonance imaging (fMRI) and correspond to those networks of brain regions that are activated during cognitive tasks, suggesting that these intrinsic networks are an important neurovascular substrate supporting cognitive functions^2^. High within-network connectivity but low between-network connectivity, called system segregation, is a key global feature associated with higher cognitive performance across the life-span^3, 4^. During aging and disease, system segregation decreases^3, 5–9^ (for review see^10, 11^), resulting in poorer cognitive functions^3, 5, 9, 12–14, 15^ (for review see^4^). In contrast, relatively preserved system segregation enhances cognitive resilience to brain alterations in older adults^16^. System segregation is thus a feature of global functional brain organisation that is crucial to sustain cognitive abilities throughout the adult life-span. However, little is known about factors that modulate system segregation^3^ and thus may yield targets for developing interventions to enhance cognitive resilience to disease.

The human brain connectome is under significant genetic control^17–20 21^. Especially higher-order features such as resting-state fMRI assessed system segregation showed high heritability ranging between 38% and 59% in twin studies^19^. Genome-wide association studies (GWAS) have provided tremendous progress in uncovering the genetic variants and potential biological pathways underlying neuroimaging phenotypes including connectivity^21^. Yet, the genetic architecture that supports system segregation of the healthy connectome is unknown. Here, we adopt a GWAS approach to identify genetic variants and associated pathways of system segregation in cognitively normal subjects spanning the adult life-span.

For our analyses, we draw on a unique set of neuroimaging and genetic data from two large-scale population-based studies: the UK Biobank^22^ and the Rotterdam Study^23^. We first performed a GWAS of system segregation complemented by pathway and tissue enrichment analysis of the associated genes based on our discovery sample (UK Biobank participants, N=16,635, 41-81 years) followed by a targeted replication analysis in the Rotterdam Study (N=2,414, 52-90 years,). Potential causal effects of the discovered pathways were subsequently assessed by Mendelian randomization (MR) analyses. GWAS in the UK Biobank identified 536 genome-wide significant single-nucleotide polymorphisms (SNPs) mapped to nine independent loci and 66 genes related to system segregation. We found an enrichment of vascular functions on the SNP and gene level. Genetic correlation analysis across the seven major modifiable cardiovascular risk factors – the so-called Life’s Simple 7 (LS7)^24^ revealed a shared genetic background between systolic blood pressure and system segregation. Our MR analysis in the UK Biobank dataset demonstrated a potentially causal link between higher blood pressure and lower system segregation, which was consistent with the observational associations found in the UK Biobank and Rotterdam Study samples. A separate MR analysis confirmed a favourable influence of higher system segregation on cognitive performance. Together, the current findings expanded our genetic understanding of a global feature of brain network organization that supports cognitive performance and further revealed high blood pressure as a potentially modifiable risk factor. Implications arise for blood pressure treatment, particularly for blood pressure-lowering trials, where system segregation could be considered as an intermediate imaging marker to detect treatment effects before cognitive decline and dementia occur.

## Results

Overview of the UK Biobank and Rotterdam Study designs and flowcharts illustrating the selection of participants can be found in Supplementary Fig. 1. Details of all included sub-samples are presented in Supplementary Table 1. Sample characteristics of the main samples (used for GWAS and polygenic prediction of system segregation) are presented in Table 1. Mean age was 63.14 [range: 45-81] years in the UK Biobank sample and 67.22 [range: 52-90] years in the Rotterdam Study sample. A bit over half of the participants were women (UK Biobank: 52.9%, Rotterdam Study: 53.5%). The group-average functional brain connectome as well as the average connectome for participants with highest versus lowest system segregation scores are illustrated in Fig. 1.

**Fig. 1.**
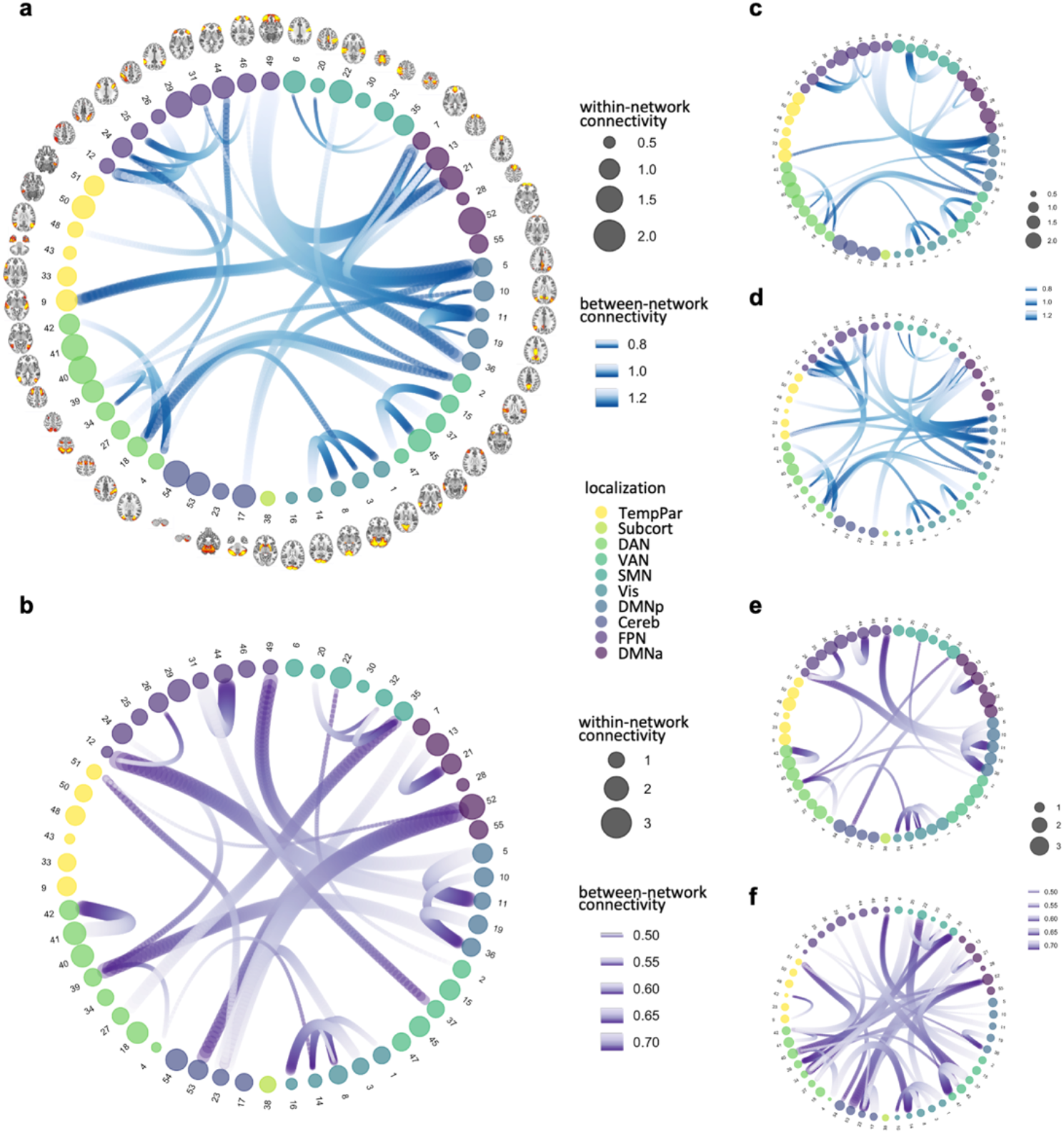
Visualization of the sample-average functional brain connectome and comparisons between participants with highest versus lowest system segregation. Average functional connectivity within and between 55 networks are displayed in **a** for UK Biobank and in **b** for Rotterdam Study participants. The 55 networks and associated spatial maps (axial view) had been derived from high-dimensional ICA performed in the UK Biobank sample. Within-network connectivity is illustrated as the group-averaged standard deviation (amplitude) of each network. Larger spheres represent stronger within-network connectivity. Between-network connectivity is illustrated as the group-averaged r-to-z-transformed correlation coefficients thresholded at 2/3 of the full range. Thicker bundles represent stronger between-network connectivity. The functional connectome of the five percent of participants with highest system segregation is displayed in **c** for the UK Biobank and in **e** for the Rotterdam Study as well as for the five percent of participants with lowest system segregation in **d** for the UK Biobank and in **f** for the Rotterdam Study. Cereb cerebellar network, DAN dorsal attention network, DMNa default mode network anterior, DMNp default mode network posterior, FPN frontoparietal network, SMN somatomotor network, Subcort subcortical network, TempPar temporoparietal network, VAN ventral attention network, Vis visual network

**Table 1.**
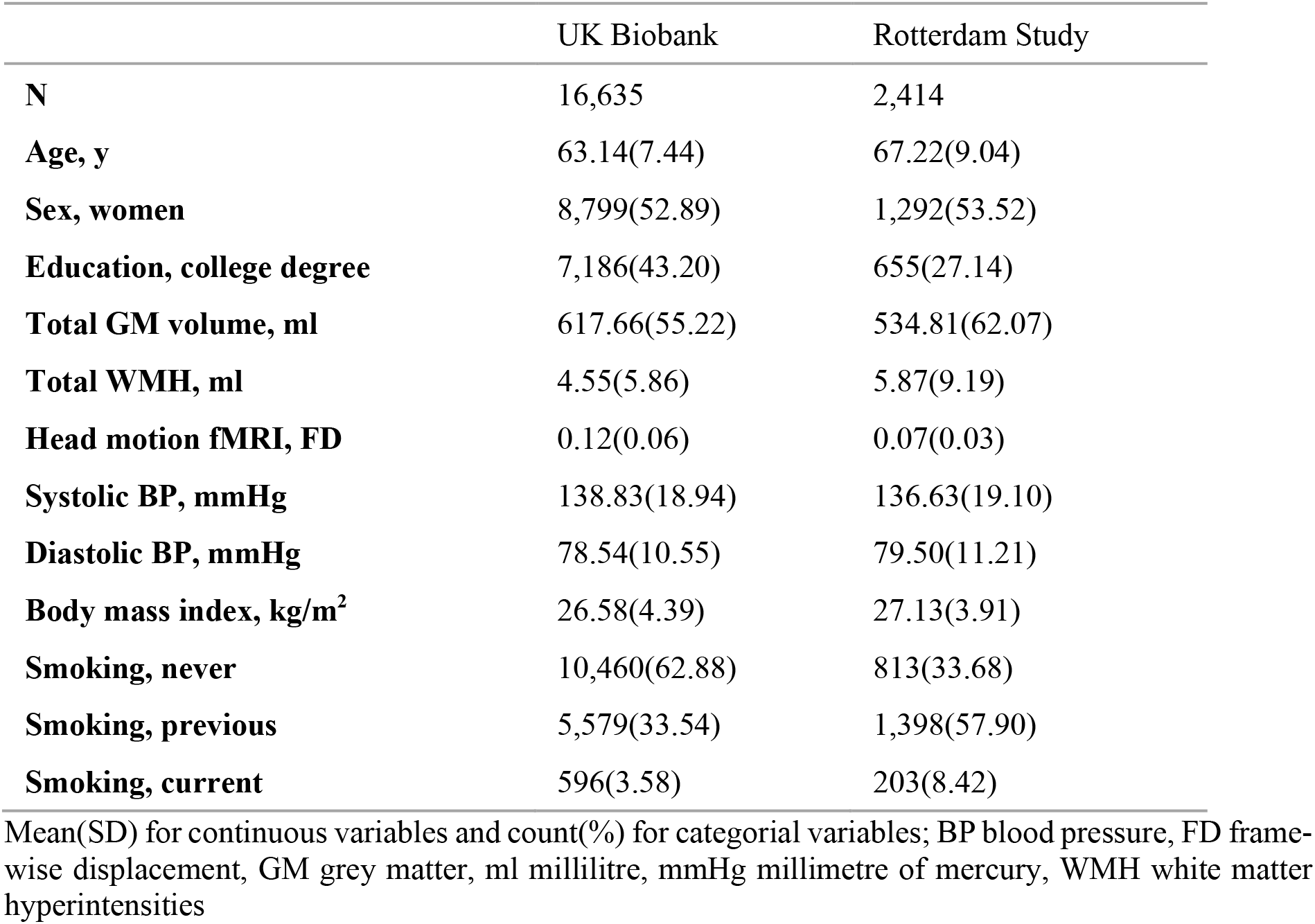
Sample characteristics

### Significant single-variant associations with system segregation

GWAS results of system segregation performed in 16,635 UK Biobank participants are displayed in Fig. 2. A total of 536 SNPs reached genome-wide significance (P < 5 × 10^-8^; Supplementary Data 1). We identified 53 independent significant SNPs (r^2^ < 0.6) and 12 lead SNPs (r^2^ < 0.1) which are associated with system segregation (respectively marked in blue and green on Manhattan plot in Fig. 2a). Based on all lead SNPs, nine independent genetic loci were detected, after merging regions < 250 KB apart into a single locus (Supplementary Fig. 2 and Supplementary Table 2).

**Fig. 2.**
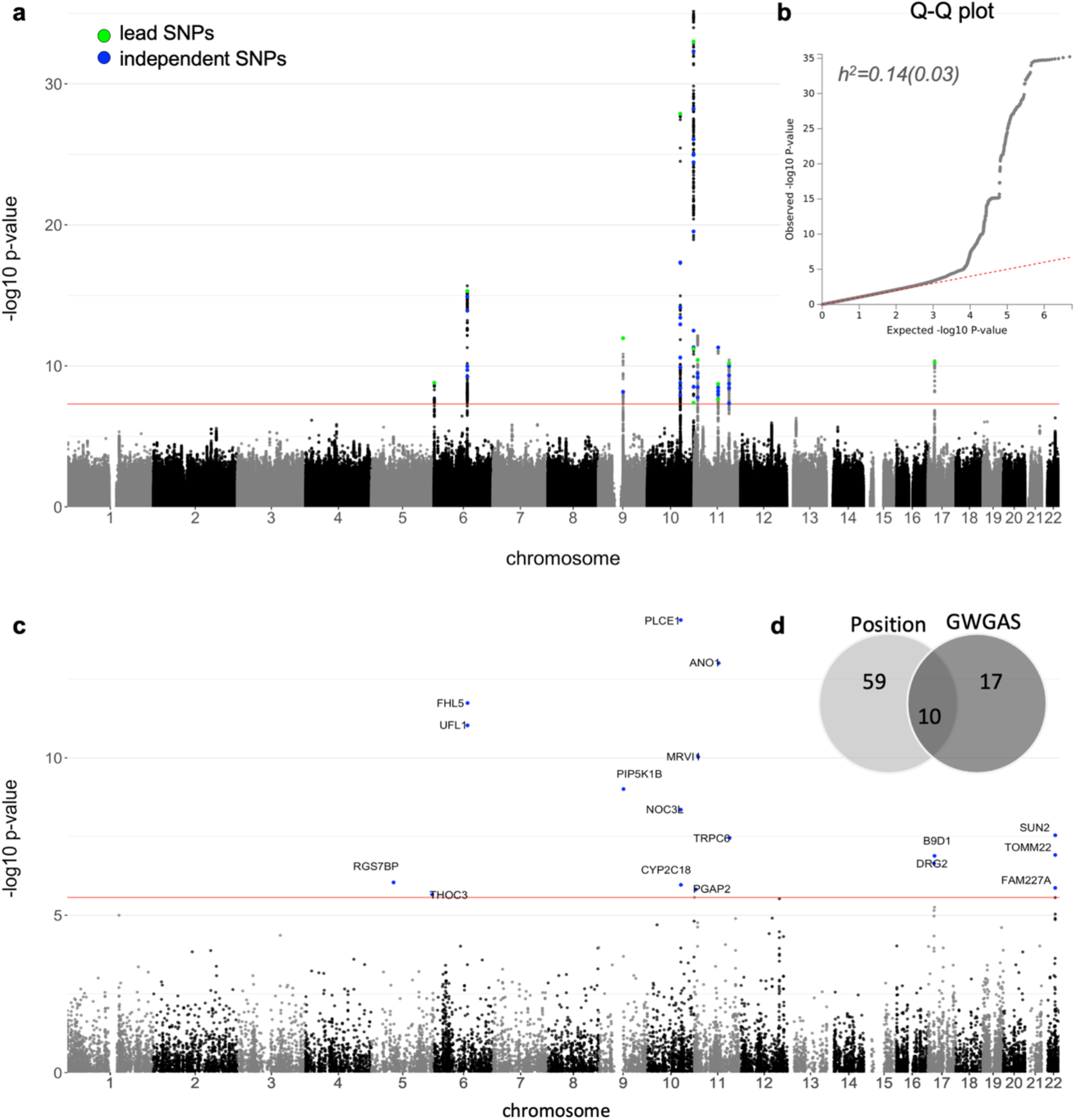
Genome-wide association analysis of system segregation in UK Biobank (N=16,635) **a** Negative log10-transformed P-values for each SNP (y-axis) are plotted by chromosomal position (x-axis). The red lines represent the thresholds for genome-wide, statistically significant associations (P = 5 × 10^-8^). Blue and green dots represent the independent significant (N=53) and lead SNPs (N=12) respectively among the 536 genome-wide significant SNPs (listed in Supplementary Data 1). **b** The quantile-quantile plot shows observed (y-axis) versus expected (x-axis) SNP P-values. The h^2^ value denotes SNP-based heritability computed by LD score regression. **c** Results of gene-based genome-wide association analysis (GWGAS) as the negative log10-transformed P-values for each gene (y-axis) plotted by chromosomal position (x-axis). The red line marks the genome-wide significance threshold at 2.73 × 10^-6^ (i.e. 0.05/number of genes). The 17 significantly associated genes are labeled. **d** Venn diagram shows overlap between 66 unique genes implicated by position mapping with FUMA or GWGAS.

The quantile-quantile (Q-Q) plot of all SNPs shows modest inflation (λ_GC_=1.04; Fig. 2b). We performed linkage disequilibrium score regression (LDSC)^25^ to quantify the proportion of inflation in the mean χ2 that was due to confounding biases. The low LDSC intercept of 1.01 indicates a small contribution of bias, and that the mean χ2 of 1.06 results mostly from polygenicity and not from other sources of bias such as population stratification. SNP-based narrow-sense heritability was estimated at h^2^ = 0.144 (standard error (SE) = 0.03).

Detailed functional annotation of all 536 significant SNPs as implemented in FUMA^26^ can be found in Supplementary Data 1. Focusing on the 12 lead SNPs (Supplementary Table 3), we found 10 SNPs in intronic or intergenic regions, yet rs1133400 and rs227422 were exonic non-synonymous SNPs and hence in coding regions. Combined Annotation Dependent Depletion (CADD)^27^ scores > 17 indicated that both SNPs are likely deleterious. The rs1133400 SNPs is a missense variant (MAF = 0.49, arginine-proline substitution) of the phospholipase C epsilon 1 (*PLCE1*) gene. G allele carrier status is associated with lower system segregation, while other recent studies in the UK Biobank have found this variant to be related with higher body fat percentage^21, 28^ and higher blood pressure^29^. The rs227422 SNP is a missense variant (MAF = 0.21, lysine-arginine subsitution) of the inositol polyphosphate-5-phosphatase A (*INPP5A*) gene, with the G allele being linked to lower system segregation. *INPP5A* controls intracellular calcium signalling^30^. Interestingly, two other lead SNPs, rs138004790 and rs9645539, with likely regulatory functionality (Regulome DB^31^ score ≤ 3) are CTCF-binding sites flanking *INPP5A*.

### Out-of-sample polygenic prediction of system segregation

Next, we estimated how much variance in system segregation measured in the independent Rotterdam Study can be explained by the current GWAS results by means of a polygenic risk score (PRS) which we computed via PRSice2^32^. In brief, different PRSs based on GWAS summary statistics were computed, iteratively included SNPs across different P-value thresholds (P < 5 × 10^-8^ – 0.5) to achieve best model fit. Strongest prediction of system segregation was accomplished when only the top 13 GWAS SNPs were considered, i.e. only SNPs with P < 1 × 10^-7^ (Supplementary Table 4). This PRS explained 1.4% of variance in system segregation in the Rotterdam Study sample (Supplementary Fig. 3a). The association between PRS and system segregation remained significant after accounting for age, age^2^, sex, education, grey matter (GM) volume, white matter hyperintensities (WMH) volume, motion during fMRI and signal-to-artefact ratio as covariates in the statistical model (standardized beta [beta_SD_] = 0.110, SE = 0.02, P < 0.001, N = 2,414, Supplementary Fig. 3b).

### Gene-based associations with system segregation

We used two independent strategies to gain insights into which genes are linked to system segregation. First, positional gene-mapping via FUMA was performed to map individual significant SNPs in the risk loci within a 10kb window to 59 genes (see Supplementary Table 2). Second, genome-wide gene-based association study (GWGAS) using MAGMA^33^ identified 17 genes to be associated by system segregation at a Bonferroni-corrected significance threshold (P < 2.72 × 10^-6^; Fig. 2c, Supplementary Data 2). MAGMA estimates aggregate associations on the basis of all SNPs in a gene. The strongest gene-based associations with system segregation were found for the *PLCE1* (P < 4.15 × 10^-15^) and anoctamin 1 (*ANO1*; P < 9.78 × 10^-14^) genes. Both genes are involved in signal transduction, either by regulating phospholipase C-activating G-protein coupled receptor signalling or by *AOV1* encoding calcium-activated chloride channels. Positional gene-mapping and GWGAS identified 10 overlapping genes (*ANO1, B9D1, CYP2C18, FHL5, MRVI1, NOC3L, PIP5K1B, PLCE1, TRPC6, UFL1*) resulting in 59+17–10=66 unique genes potentially associated with system segregation (Fig. 2d) which were further explored via gene expression and gene set analyses. In gene expression analysis, we assessed whether the 66 genes of interest are overexpressed in any of the 54 different tissue types included in the GTEx v8 database at a Bonferroni-corrected significance threshold. We found a negative association (downregulation) with gene expression across multiple brain areas, while a positive association (upregulation) with gene expression in cardiovascular tissue (coronary, tibial and aorta artery) and adipose tissue was observed (all P < 9.26 × 10^-4^; Fig. 3a).

**Fig. 3.**
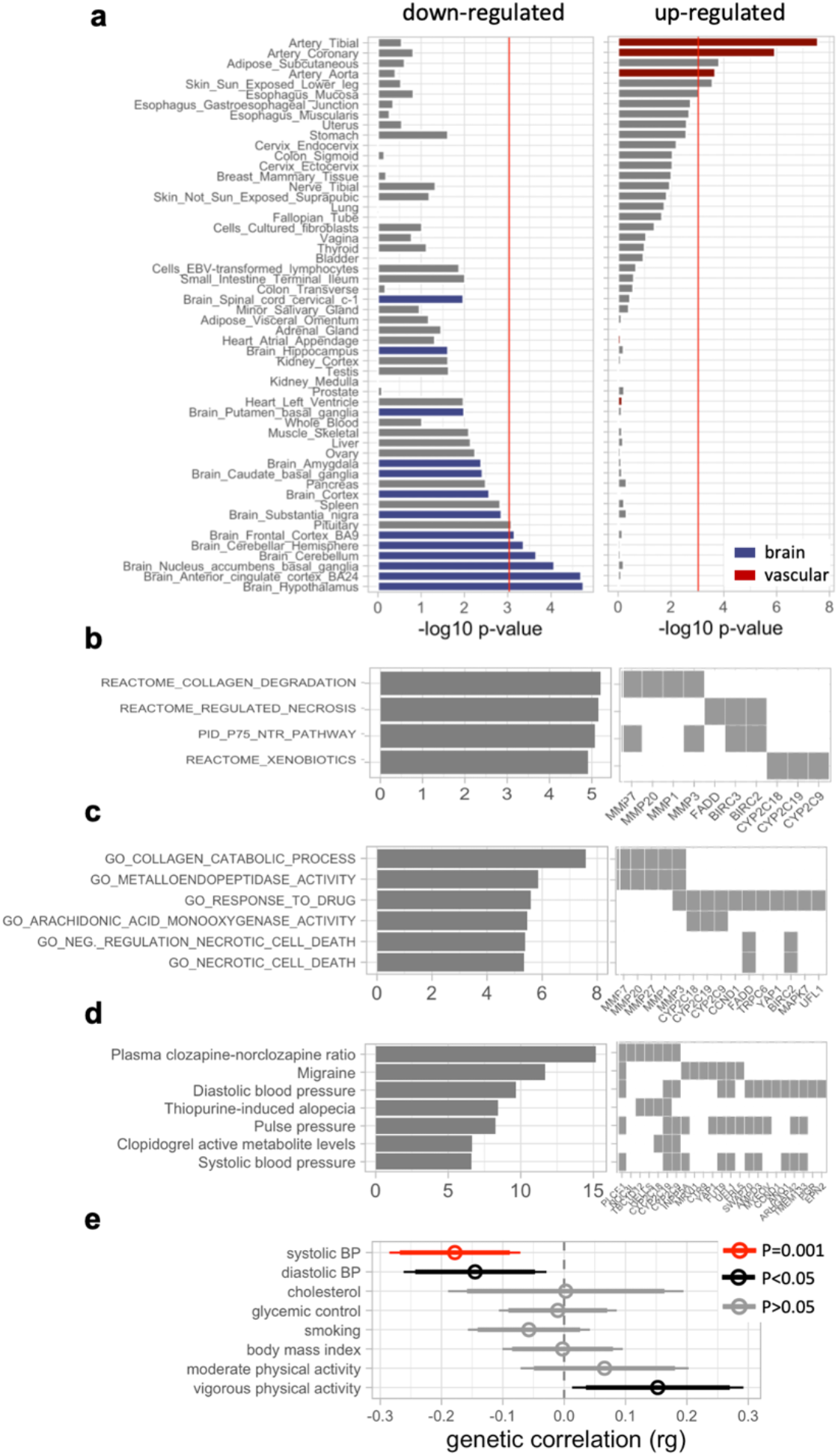
Post-GWAS analyses results of system segregation in the UK Biobank (N=16,635) **a** Results of differential gene expression analysis are plotted as the negative log10-transformed P values for 54 GTEx tissue types. The red line marks the Bonferroni-corrected significance threshold at 9.26 × 10^-4^ (i.e. 0.05/number of tissues). FUMA differentiates upregulated and downregulated genes dependent on the sign of the t-test. We color-coded brain tissue in blue and vascular tissue in red. Results of gene set analysis are displayed in **b** for canonical (N=2,868) and in **c** for GO (N=7,573) pathways as well as in **d** for genes reported in the GWAS catalogue. Only associations at a Bonferroni-corrected significance threshold (0.05/number of pathways in each category) are shown. Bars indicate the negative log10-transformed P-value, while the squares show the inputted genes that are part of the enriched gene sets. All analyses were performed using FUMA^26^. **e** Results of LD score regression analyses are shown testing for a genetic correlation between system segregation and seven cardiovascular risk factors (blood pressure, blood cholesterol, glycaemic control, smoking status, body mass index, physical activity and diet) measured by 27 variables. Statistical results for variables related to diet are in Supplementary Table 8 (all P > 0.05). Point estimates for genetic correlations (rg) and 95% confidence intervals are shown. Bonferroni-corrected P-values are color-coded in red.

Next, we performed gene set analysis in FUMA testing enrichment of canonical pathways (N=2,868) and GO gene sets (N=7,573). Results are reported at a stringent Bonferroni-corrected threshold taking the number of tests performed per gene set category into account. Four canonical pathways (all P < 1.74 × 10^-5^, Fig. 3b, and Supplementary Table 5a) and six GO gene sets (all P < 4.87 × 10^-6^; Fig. 3c, and Supplementary Table 5b) were significantly associated with the 66 genes of interest. Several pathways point towards molecular processes important for blood vessel structure and remodelling, including collagen processing and matrix metalloproteinases, and which had been shown to play a role in small vessel disease and stroke^34^. Of note is the association with arachidonic acid activity which among others is involved in regulating blood pressure as well as in the coupling between the metabolic activity of neurons and regional blood flow in the brain^35^. Additionally, we conducted gene set analysis for all gene set reported in the GWAS catalogue (N = 2,195). Seven significant associations were found at a Bonferroni-corrected significance threshold (P < 2.28 × 10^-5^). These results can be divided into three main groups including associations with genes related to drug effectiveness, migraine and blood pressure (Fig. 3d, and Supplementary Table 6).

### Genetic correlations between system segregation and cardiovascular risk factors

Since we found a distinctive upregulation of the genes of interest in vascular tissue as well as enrichment of gene sets related to vascular functions, we evaluated the shared genetic environment between system segregation and cardiovascular risk factors using LD score regression^25^. For this purpose, we utilized previously calculated LD summary scores of the seven main cardiovascular risk factors (Life’s Simple 7, LS7) defined by the American Heart Association^24^, i.e. blood pressure, blood cholesterol, glycaemic status, smoking, body mass index, physical activity and diet (accessible via https://nealelab.github.io/UKBiobankB_ldsc/downloads.html; see Supplementary Table 7a for UK Biobank data fields). We observed a negative correlation with systolic blood pressure (rg = -0.178, SE=0.055, P = 0.001) at a Bonferroni-corrected significance threshold. We also found a negative correlation with diastolic blood pressure (rg = -0.145, SE = 0.059, P = 0.014) and a positive correlation with physical activity (rg = 0.153, SE = 0.071, P = 0.033), yet both results did not pass multiple test correction (Fig. 3e, and Supplementary Table 8).

### Lower blood pressure is associated with higher system segregation

Next, we estimated the relationship between LS7 cardiovascular risk factors and system segregation using observational data of all UK Biobank participants included in the GWAS (see Methods). Cardiovascular health scores were coded as poor=0, intermediate=1, and optimal=2 according to the American Heart Association recommendations and considering pharmacological treatment (for cut-off definitions see Supplementary Table 7b)^24^. All seven cardiovascular traits were measured at baseline (on average 7.6 [range: 4-12] years prior to brain imaging) and five traits were measured at the time of brain imaging, not including blood cholesterol and glycaemic status (blood was drawn only at baseline). All statistical models were adjusted for age, age^2^, sex, education, GM volume, WMH, head motion during fMRI, assessment centre (only for UK Biobank) and signal-to-artefact ratio (only for Rotterdam Study). Consistent with the genetic correlation analysis, we found a significant association between blood pressure and system segregation, where an optimal blood pressure score (i.e. SBP <120 mm Hg and DBP <80 mm Hg untreated) was related to higher system segregation (poor versus optimal score at baseline visit: beta = 0.142, SE = 0.020, P < 0.001, N = 16,635, or at imaging visit: beta = 0.159, SE = 0.021, P < 0.001, N = 16,635; Fig. 4a). A trend-level association was found between optimal blood cholesterol levels (LDL-C < 130 mg/dl) and higher system segregation which did, however, not survive Bonferroni-correction. No other significant association was found (all P > 0.05; Supplementary Table 9). The reported findings remained consistent when LS7 risk factors were considered as continuous predictors (systolic blood pressure at imaging visit: beta_SD_ = -0.059, SE = 0.007, P<0.001), Supplementary Fig. 4a) or after re-running the analysis in a non-imputed dataset (systolic blood pressure at imaging visit: beta_SD_ = 0.044, SE = 0.004, P < 0.001, Supplementary Fig. 4b).

**Fig. 4.**
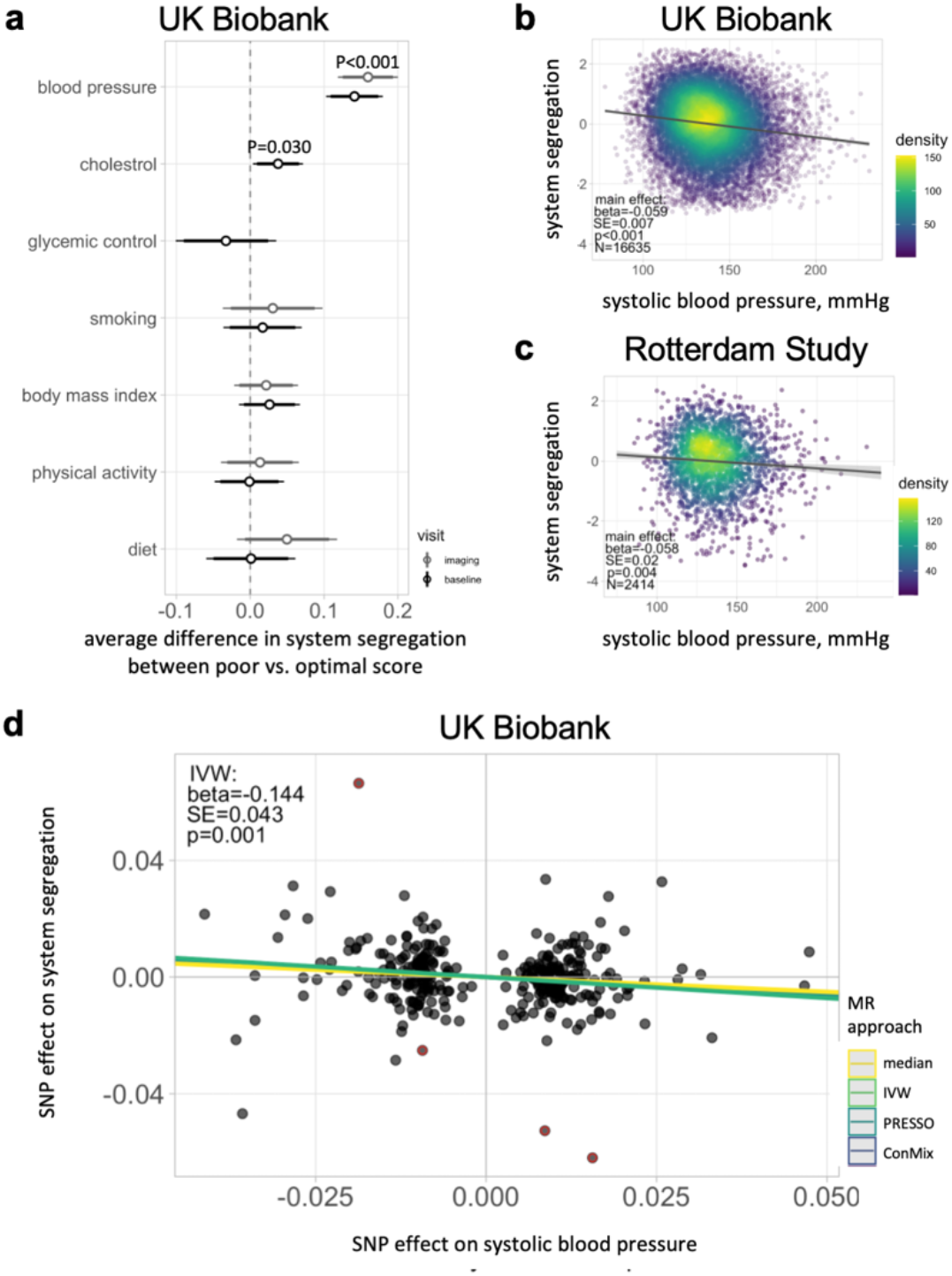
Association between cardiovascular risk factors and system segregation. **a** Whisker plot displays associations between Life’s Simple 7 (LS7) cardiovascular risk factors (all classified on a 3-point scale according to AHA criteria) and system segregation (z-score) in UK Biobank participants. All scores were assessed at baseline and non-blood-based measurements were re-assessed at the brain imaging visit. Beta coefficients together with 95% confidence intervals are shown. Scatter plots show the main effect of continuous values of systolic blood pressure (mmHg) on system segregation (z-score) in **b** for UK Biobank participants (at imaging visit) and in **c** for Rotterdam Study participants (replication analysis). All statistical results are derived from multiple regression models including age, age^2^, sex, education, grey matter volume, white matter hyperintensity volume, head motion during fMRI and assessment centre (only UK Biobank) or signal-to-artefact ratio (only Rotterdam Study) as covariates. Standardized beta values and standard errors (SE) are reported. Linear model fits are displayed together with 95% confidence intervals. **d** The scatter plot displays the results of two-sample Mendelian randomization analysis in the UK Biobank estimating the causal effect of systolic blood pressure (x-axis) on system segregation (y-axis). The genetic instruments of blood pressure (N=312) were derived from GWAS results of a non-overlapping UK Biobank sample without brain imaging (N=364,061). Statistics are reported for the main analysis using random-effects IVW (green line). Results of alternative MR approaches are additionally displayed. ConMix contamination mixture method, IVW inverse-variance weighted method, PRESSO Pleiotropy RESidual Sum and Outlier method

The replication analysis in the Rotterdam Study sample confirmed a significant association between higher blood pressure (continuous predictor) and lower system segregation (beta_SD_ =- 0.058, SE = 0.020, P=0.004, N = 2,414, Fig. 4c).

### Mendelian randomization association between genetic predisposition to higher blood pressure and lower system segregation

In two-sample MR analyses, we estimated the causal effect of blood pressure on system segregation. 312 independent genetic instruments of systolic blood pressure were identified based on GWAS output (all SNPs with P < 5 × 10^-8^ and r^2^ ≤ 0.001; Supplementary Data 3) from a non-overlapping sample of UK Biobank participants for whom fMRI was not available (N=364,061). Random-effects inverse variance weighted (IVW) MR analysis yielded a significant negative association between genetic variants of blood pressure and system segregation (beta_SD_ = -0.144, SE = 0.043, P = 0.001, Fig. 4d). Test of heterogeneity was significant which indicates that one or more variants may be pleiotropic (Cochran’s Q, P < 0.001). Directed pleiotropy has unlikely biased IVW estimates, since the MR Egger intercept was not significantly different form zero (Egger intercept: 0.001, SE = 0.001, P = 0.332) and the funnel plot was symmetrical (Supplementary Fig. 5a). Alternative MR approaches, which are more robust in the presence of pleiotropy, were performed (Supplementary Method 2). MR-PRESSO identified four outliers (rs11153071, rs11187838, rs80226362, rs9869147). The outlier-corrected estimate from MR-PRESSO was comparable to the IVW result (beta_SD_ = - 0.119, SE = 0.039, P = 0.002) and also the contamination mixture approach (beta_SD_ = -0.118, SE = 0.105, P = 0.014) and weighted median approach (beta_SD_ = -0.101, SE = 0.052, P = 0.052) yielded relatively comparable associations (Fig. 4d, Supplementary Table 10). Leave-one-out sensitivity analysis showed consistent results indicating that the result is unlikely biased by outliers (Supplementary Fig. 5b).

### Higher system segregation is associated with better cognitive performance

Based on previous studies reporting higher system segregation to be related with better cognitive performance in elderly participants, we determined age-dependent and -independent effects of system segregation on cognition. Out of the 19,822 UK Biobank participants for whom fMRI data was available at the time of download [March 2019], a subsample of 7,342 participants completed the enhanced psychometric test battery at the brain imaging visit. Of the 7,342 participants 2,113 underwent a follow-up cognitive assessment on average 2.23 [range 2-3] years after the imaging visit. Cognitive performance was defined by a factor score computed based on six psychometric tests which covered memory, executive functions, as well as verbal and non-verbal reasoning skills (see Methods). All models were adjusted for age, age^2^, sex, education, GM volume, WMH volume and assessment centre (only for UK Biobank). We found higher system segregation to be associated with better cognition (beta_SD_ = 0.031, SE = 0.011, P = 0.005, N = 7,342; Fig. 5a). No system segregation × age interaction was observed (beta_SD_=-0.012, SE=0.011, P=0.288, N=7,342). In the subsample with follow-up cognitive assessment available, mixed effects regression analysis revealed that higher system segregation was associated with slower rates of cognitive decline (beta_SD_ = 0.035, SE = 0.015, P = 0.018, N = 2,113, Fig. 5b).

**Fig. 6.**
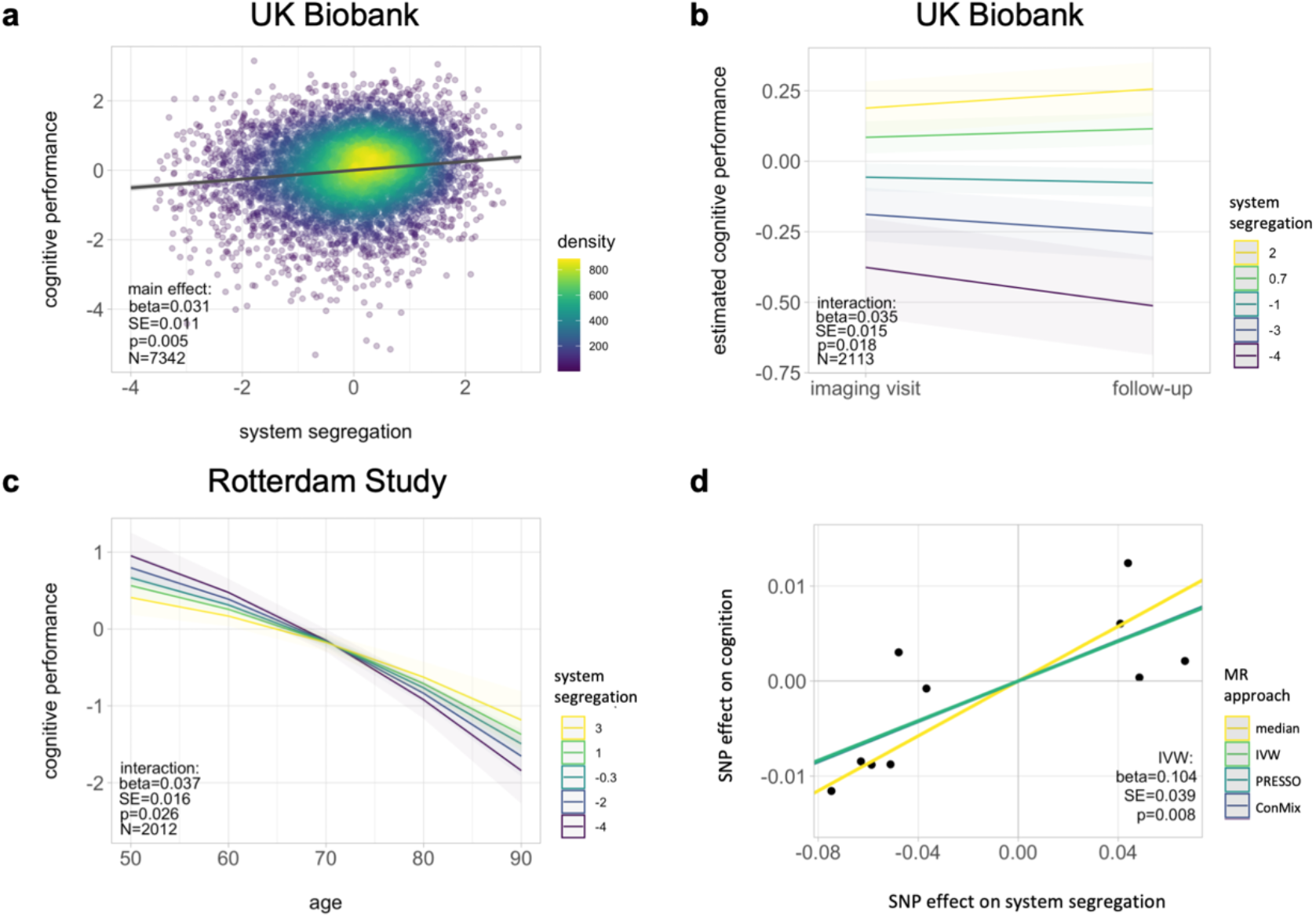
Association between system segregation and cognitive performance. **a** Scatter plot shows the main effect of system segregation (z-value) on cognitive performance (z-value), i.e. a factor score derived from six tests examining distinct cognitive domains, in the UK Biobank sample. **b** Line plot displays the interaction between study visit (x-axis) and system segregation (color-coded) on mean predicted cognitive performance (y-axis) in UK Biobank participants with cognitive follow-up assessment. Note that we plotted the fitted value of cognitive performance, when the random effect for participant is held constant. **c** Results of the cross-sectional replication analysis in the Rotterdam Study sample are illustrated as the interaction between age (x-axis) and system segregation (color-coded) on cognitive performance (y-axis), i.e. a factor score derived from five tests examining distinct cognitive domains. Statistical results are derived from linear regression models (for a, c) or a linear mixed effects model (for b) including age, age^2^, sex, education, grey matter volume, white matter hyperintensity volume, assessment centre (only UK Biobank) and random intercept for participant (only mixed-effects model) as covariates. Linear model fits are indicated together with 95% confidence intervals. **d** Scatter plot displays the association between genetic predisposition of system segregation (x-axis) and cognition (y-axis) computed by MR analysis in the UK Biobank sample. Genetic instruments for system segregation (N=10) were derived from the current GWAS results. The genetic variants for cognition are based on GWAS results of a non-overlapping UK Biobank sample with cognitive assessment, but no fMRI available. Statistics are reported for the main analysis using random-effects IVW (green line). Results of alternative MR approaches are additionally shown (for full statistical results see Supplementary Table 10). In all plots, we reported standardized beta values. IVW inverse-variance weighted method, PRESSO Pleiotropy RESidual Sum and Outlier method, ConMix contamination mixture method.

Out of the 3,288 Rotterdam Study participants with fMRI available [December 2020], 2,012 participants completed cognitive assessment at the brain imaging visit. Follow-up cognitive assessment was not available. Cognitive performance was defined by a factor score computed based on five neuropsychological tests which covered memory, executive functions, as well as verbal fluency and motor skills (see Methods). We found no main effect of system segregation on cognitive performance (beta_SD_ = -0.018, SE = 0.015, P = 0.335, N = 2,012). However, a significant system segregation × age interaction was observed (beta_SD_ = 0.037, SE = 0.016, P = 0.026, N=2,012, Fig. 5c) indicating that higher system segregation was associated with better cognition in the oldest but not in the younger participants. This discrepancy between the two samples could be due to differences in study design, sample size or neuropsychological/fMRI assessment.

### Mendelian randomization association between genetic predisposition to higher system segregation and better cognition

To explore the causal effect of system segregation on cognition, two-sample MR analysis was carried out. Ten genetic instruments of system segregation were derived from the current GWAS (Supplementary Data 4). GWAS summary results for cognitive performance was derived from a non-overlapping UK Biobank sample (N=10,558) for which cognitive assessment, but not fMRI, was available. Using the IVW approach, we found a significant association between higher genetic disposition of system segregation and cognitive performance (beta_SD_ = 0.104, SE = 0.039, P = 0.008, Fig. 5d). Pleiotropy unlikely influenced the results based on a non-significant test of heterogeneity (Cochran’s Q, P = 0.827). A comparable positive association between system segregation and cognition was also found when using alternative MR methods (Supplementary Table 10). MR-PRESSO identified no outliers.

## Discussion

We discovered 12 lead variants, 9 independent risk loci and 66 genes that were associated with differences in the brain’s segregated network architecture. Many of the variants related to system segregation point towards vascular functions. LD score regression indicated a specific link between higher systolic blood pressure and lower system segregation, which was confirmed in observational and MR analyses, and replicated in an independent sample. In addition, we extended earlier work by MR evidence demonstrating that lower system segregation was related to poorer cognition.

The top SNPs discovered in the GWAS linked to system segregation, and which are likely regulatory or deleterious, were related to the *INPP5A* (rs138004790, rs9645539, rs1133400) and *PLCE1* (rs2274224) genes. *PLCE1* was also the top hit of the gene-based analysis. Both genes play a key role in calcium signalling by generating (*PLCE1*) or modulating (*INPP5A*) the second messenger molecule inositol 1,4,5-trisphosphate (IP_3_) which in turn binds to the IP_3_ receptor, stimulating the transient release of calcium from the endoplasmic reticulum. Since inositol calcium signalling is such a fundamental cellular process, it likely contributes to multiple neurobiological mechanisms underlying functional connectivity. This notion is supported by two previous UK Biobank GWAS highlighting the rs2274224 SNP and a LD proxy of rs9645539 (i.e. rs11596664, r^2^=0.99) as two of the top hits for frontal cortex connectivity^21, 36^. In another UK Biobank study, the *INPP5A* and *PLCE1* regions were associated with the proportion of brain age explained by changes in functional connectivity^37^. Another strong genetic signal came from a locus on chromosome six encompassing the *FHL5* and *UFL1* genes. By regulating smooth muscle cell contraction, multiple of the SNPs we identified in this locus have previously been implicated in blood pressure (rs11153018, rs3798293)^38^, cerebral blood flow (rs2971609)^39^ and migraine (rs7775721, rs10786156)^40^. The same mechanisms may explain the link to system segregation, and more generally to functional connectivity of many brain networks as shown earlier (rs3849198, rs11153070, rs12204342, rs6921291)^21, 36^.

Four of the nine loci (#1,3,6,9) we discovered in the current study were not found in the two previous GWAS on functional connectivity,^21, 36^ suggesting a more specific contribution to system segregation. An interesting novel locus was detected on chromosome six with the lead SNP rs7766042 being a regulatory variant of the *FOXF2* gene. This SNP is a known risk variant for ischemic stroke (OR: 2.96) and small artery occlusion (OR: 3.35)^41^. Inactivation of *FOXF2* in animals resulted in cerebrovascular defects and a breakdown of the blood-brain barrier due to *FOXF2* crucial role in the differentiation of brain pericytes^42^.

We found another novel locus on chromosome seventeen implicating the Epsin 2 (*EPN2*) gene which is enriched in the brain and involved in clathrin-mediated endocytosis. Mice conditionally lacking epsins display embryonic lethality because of vascular defects suggesting that epsins are required for normal development of the vasculature^43^. A recent GWAS in the UK Biobank related an LD proxy of the lead SNP rs72639204 (i.e. rs6587216, r^2^=0.97) to cerebral white matter hyperintensity volume^44^. Structural and functional connectivity are highly interrelated^45^ and reduced integrity of the former seems a plausible mechanism for compromising system segregation. In addition, these findings may suggest that changes in system segregation could be a novel neuroimaging marker of cerebrovascular disease, a hypothesis which should be tested in future research.

This potential link to vascular pathways was affirmed in our post-GWAS analysis. Besides the SNP and locus-based associations reported above, the identified genes were significantly overexpressed in vascular tissue (particularly in arteries) and enriched for pathways critical for tissue remodelling and neurovascular coupling. These findings appear plausible considering the neurons and vascular cells have a close developmental, structural, and functional relationship^46^. Neurovascular coupling, i.e. the increased local blood flow and tissue oxygenation in response to neuronal activity, is a fundamental brain property that supports local neuronal activity through oxygen and metabolic supply. The vascular responsivity has thus a vital function in sustaining neuronal activation during cognitive processes. Resting-state fMRI utilizes neurovascular coupling to infer functional connectivity at the scale of ultra-slow vasomotor fluctuations of brain vessels that oscillate at 0.1 Hz matching that of ultra-slow rhythmic neural oscillations^47^. Experimentally induced changes in vascular reactivity after inhalation of CO_2_-enriched air propagate at the network level, mirroring the spatial distribution of fMRI detected functional large-scale networks such as the default model network^48^. Therefore, one possibility is that disturbances of vascular reactivity such as related to hypertension may alter functional network connectivity at a global scale including network segregation. However, direct evidence for changes in oscillatory vascular activity altering functional connectivity is missing to date. The current results provide genetic evidence for systemic vascular alterations to be associated with system segregation.

Of the seven main cardiovascular risk factors, we showed that higher blood pressure was distinctively related to lower system segregation across genetic and observational correlation analyses and independent samples. Our MR analysis in the UK Biobank provides additional evidence for a causal role of the effect of higher blood pressure on lower system segregation. Recent data in non-demented adults at risk of Alzheimer’s disease consistently showed that high blood pressure, but not high cholesterol, was linked to a global reduction of network connectivity assessed over time^49^. Although the exact mechanisms of how hypertension affects system segregation are insufficiently understood, there are many biologically plausible pathways. At the molecular level, hypertension increases oxidative stress due to excess production of vascular reactive oxygen species (ROS)^50^. Oxidative and mechanic stress have been linked to profound cerebrovascular changes (including atherosclerosis, arterial stiffening, impaired cerebrovascular autoregulation and reactivity; for reviews see^51, 52^), for some of which we found evidence in the pathway analysis. Remodelling and stiffening of blood vessels should protect downstream microvessels from the stress of chronic hypertension but which become maladaptive^53^. Our pathway analysis consistently showed enrichment for collagen and matrix metalloproteinases gensets which play an essential role in angiogenesis and stiffening of blood vessels^53^. Increased production of ROS has been also related to reduced endothelial nitric oxide (NO) bioavailability, impaired NO-dependent endothelial vasodilation, ultimately compromising neurovascular coupling which is essential for neuronal activity^51^. To better understand the underlying processes between high blood pressure and system segregation, it would be interesting to investigate the relationship in conjunction with other relevant imaging modalities such as blood perfusion, arterial stiffness and calcification imaging.

By extending earlier observational analyses, our current MR approach, additionally demonstrated that higher system segregation contributes to better cognitive performance. The cross-sectional and longitudinal observational data in the UK Biobank aligns with this conclusion. In the Rotterdam Study, higher system segregation was associated with better cognitive performance only in the oldest participants, a finding which may suggest that more clearly segregated brain networks become especially relevant for individual at high risk of cognitive decline. This idea, that higher system segregation reflects one aspect of brain resilience, is supported by our previous study where we showed that higher system segregation was related to better cognition in the presence of Alzheimer’s disease pathology^16^. Alternatively, differences in the UK Biobank’s and Rotterdam Study’s design and sample size may have caused the discrepancy.

Another interesting but yet to be tested hypothesis emerging from the current data implicates that changes in system segregation due to high blood pressure may be a pathway by which poor cardiovascular health is linked to cognitive impairment and dementia^54^. In agreement with this notion are the results from a recent study in the UK Biobank which show that a history of hypertension was related to reduced hippocampus connectivity which in turn was associated with poorer memory performance^55^. Together, these results may be valueable for blood pressure-lowering randomized controlled trials which so far failed to show a clear treatment effect on dementia outcome^56^ in contrast to the beneficial effects reported across many epidemiological studies^57^. Global measures of network connectivity, such as system segregation, may be considered as complementary imaging markers to detect intermediate treatment effects before cognitive decline or dementia occur.

The current study is not without limitations. First, it is important to note that all study participants were of European ancestry and our findings should therefore be generalized only with great caution. Second, the SNP heritability estimate of the current study was significantly lower than the ones reported previously in family-based studies, but is in a comparable range as those observed by previous GWAS on functional connectivity^21, 36^. Third, the fMRI protocols and MRI scanners of the UK Biobank and Rotterdam Study are quite different and the Rotterdam Study subsample in which fMRI was acquired is relatively small. Both factors have likely reduced the statistical power of the polygenic prediction analysis. On the other hand, the association strength between higher blood pressure and lower system segregation is identical between the two cohorts, supporting the robustness of our findings.

In conclusion, the current genetic and observational results highlight the importance of blood pressure control for maintaining segregated brain networks which in turn was shown to benefit cognitive functions in the general aging population.

## Methods

### UK Biobank

Participants: The UK Biobank is an ongoing, longitudinal cohort study which aims to identify genetic and nongenetic determinants of the diseases of middle and old age^58^. Supplementary Fig. 1a provides a schematic overview of the three visits relevant for the current study. Baseline data collection including genotyping of 500,000 participants aged 40–69 years had been carried out between 2006 and 2010. From 2014 onwards, a subsample of participants was reinvited for brain imaging (including fMRI), while a third follow-up started in 2019. A flowchart of participant selection can be found in Supplementary Fig. 1b. After quality assessment performed by UK Biobank^59^, fMRI data of 19,822 participants were available and downloaded as of data release in March 2019. We additionally excluded participants for whom fMRI connectivity analysis failed (N =760) and participants with a diagnosis of dementia or stroke (N = 112). After quality assessment of participants’ genotype data, a total of 16,635 participants remained in the current main analysis (i.e. GWAS on system segregation).

Genotyping: At the baseline visit, participants were genotyped using either the UK Bileve or Axiom array. We used the imputed genetic dataset made available by UK Biobank in its version 3 release. This consists of ∼96 million genotypes imputed from the Haplotype Reference Consortium (HRC) reference panel and a merged UK10K + 1000 Genomes reference panel. In addition to the quality control carried out by UK Biobank^60^, we excluded samples with a call rate below 97%, gender mismatches, excess autosomal heterozygosity, family relations and non-white British ancestry, as well as vaiants with minor allele frequencies below 1%,.

Imaging: Details about the acquisition and processing of the UK Biobank imaging data can be found in the online documentation (http://Biobank.ctsu.ox.ac.uk/crystal/crystal/docs/brain_mri.pdf). Briefly, fMRI has been performed on a Siemens Skyra 3T with the following parameters: 32-channel RF receive head coil, TR = 753 ms, TE = 39 ms, Field-of-view (FOV) = 88 x 88 x 64, resolution = 2.4 mm^3^, 8-fold multislice acceleration and for 6:10 min (490 timepoints). Preprocessing and group-level independent component analysis (ICA) were carried out by UK Biobank via FSL packages. This preprocessing pipeline included motion correction, grand mean intensity normalization, high-pass temporal filtering, echo-planar image unwarping (incorporating T1-registration), gradient distortion correction (utilizing acquired field maps), removal of structured artifacts via FSL’s ICA+FIX and normalization to MNI space^22, 59, 61^. T1 and T2-FLAIR images were acquired during the same imaging session. We used total GM volume (data field: 25006) and total volume of WMH (data field: 25781) as global estimates of structural brain integrity.

Cognitive assessment: At the imaging visit, participants underwent the enhanced version of the UK Biobank cognitive test battery^62^ (https://Biobank.ndph.ox.ac.uk/showcase/showcase/docs/Fluidintelligence.pdf). We included the following tests: Pairs Matching Test (visual memory, data field: 399, log+1 transformed), Numeric Memory Test (working memory, data field: 4282), Symbol Digit Substitution Test (processing speed, data field: 23324), Trail Making Test part B (executive function, data field: 6350, log transformed), Fluid Intelligence Test (verbal and numeric reasoning, data field: 20016), and Matrix Pattern Completion (non-verbal reasoning, data field: 6373). Complete cognitive data was available for 19,630 participants of whom 7,347 individuals also had a successful fMRI assessment. Confirmatory factor analysis (R lavaan package, v0.6.7) was used to compute a score of general cognitive performance for each participant. All cognitive tests showed a significant intercorrelation of Pearson r-values between 0.16 and 0.48 which are comparable to previous reports^62^. Factor loadings of the single cognitive scores on the latent variable (i.e. general cognitive abilities) were moderate to high (range of beta-values: 0.30 – 0.7) and fit statistics indicated a valid model fit (adjusted goodness of fit index (AGFI) = 0.960; Supplementary Table 11a).

### Rotterdam Study

Participants: The Rotterdam Study is an ongoing, longitudinal, population-based cohort study in the well-defined Ommoord district in the city of Rotterdam in the Netherlands which aims to determine the causes of common diseases in the elderly^23^. A schematic overview of the relevant visits and selection of participants is provided in Supplementary Fig. 1c and d. The Rotterdam Study was initiated in 1990 with a total of 14,926 participants being included aged 45 years or older. Genotyping was performed in 11,502 participants at the initial visit. fMRI was acquired between 2012 and 2016 covering a total of 3,288 participants. After excluding participants with poor fMRI data quality (n=293), cortical infarcts on MRI (n=80), and with prevalent diagnosis of dementia or stroke (n=37), 2,878 participants remained. After quality assessment of participants’ genetic data, a total of 2,414 participants were included in the current main analysis (polygenic prediction of system segregation).

Genotyping: Genotyping was performed using Illumina 550K, 550K duo, or 610 quad arrays. Genotypes were imputed using the MaCH/minimac software to the 1000 Genomes phase I version 3 reference panel (∼30 million SNPs)^63^. Samples with a call rate below 97.5%, as well as gender mismatches, excess autosomal heterozygosity, duplicates or family relations, ethnic outliers, variants with call rates lower than 95.0%, failing missingness test, Hardy-Weinberg equilibrium P < 1e-06, and minor allele frequencies below 1%, were removed.

Imaging: Neuroimaging was performed on a Signa Excite II GE 1.5T with the following parameters: 8-channel head coil, TR = 2900 ms, TE = 60 ms, FOV = 64 x 64 x 31, resolution = 3.3 mm^3^, and for 7:44 min (165 timepoints). The FSL-pipeline used for fMRI preprocessing^8^ was largely comparable to the pipeline used in UK Biobank. Due to a technical issue, participants were scanned with the phase and frequency encoding direction swapped. In order to correct for any ghost artefacts, we computed a signal-to-artefact ratio defined by dividing the median intensity of any ghost artefacts outside of the brain by the median intensity of signal within the brain for each participant^8^. Signal-to-artefact ratio was considered as a covariate in the statistical analyses and scans with signal-to-artefact ratio > 0.1 were excluded. To estimate overall structural brain integrity, measures of total grey matter and WHM volume were derived from T1 and T2-FLAIR images.

Cognitive assessment: Cognitive tests were performed at the imaging visit or no more than 1 year (mean = 0.36 years) apart from fMRI^64^. The following tests were included: Delayed Recall Task (verbal memory), Verbal Fluency Test (crystallized abilities), Letter-Digit Substitution (processing speed), Stroop Task (executive function), Purdue Pegboard Test (motor skills). Complete neuropsychological data was available for 10,164 participants of whom 2,012 also had fMRI. All tests showed a significant intercorrelation with Pearson r-values ranging from 0.29 to 0.52. Identical to the UK Biobank, a score of general cognitive performance was computed. Factor loadings were moderate to high with standard coefficients between 0.54 and 0.79. Fit measures showed an excellent model fit (AGFI = 0.979; Supplementary Table 11b).

### Functional connectivity analysis and system segregation

We used the brain parcellation scheme generated based on the UK Biobank fMRI dataset^59^. In brief, group-level ICA was performed on the preprocessed fMRI data, using FSL’s MELODIC to estimate 100 components. Next, The UK Biobank Imaging Core classified the components into either noise (e.g. stemming from pulsation or motion) or signal components (stemming from a neuronal source). Fig. 1a illustrates the 55 signal components as an overlay on a standard brain template. A 3D rendering of each signal components can be inspected via UK Biobank’s online visualization tool (https://www.fmrib.ox.ac.uk/UKBiobankiobank/group_means/rfMRI_ICA_d100_good_nodes.html). Dual-regression was performed in both the UK Biobank and Rotterdam Study to estimate participant-specific timecourses for all 55 component which were then entered into connectivity analysis (components are for this purpose referred to as networks). First, we computed the standard deviation of the networks’ timecourses as a measure of within-network connectivity strength (data field: 25755). Second, we computed the average correlation between each network’s timecourse with every other network’s timecourse as a measure of between-network connectivity strength. For this purpose, the node-by-node cross-correlation matrix was computed for each participant and correlation coefficients were normalized by Fisher’s r-to-z-transformation (data field: 25751). Only positive ROI-to-ROI correlations were kept according to previously established protocol for computing system segregation^3^.

System segregation was then computed as the difference between participants’ connectivity strength within versus between brain networks given by the following formula:

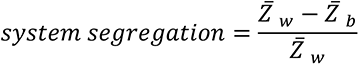

where *Z̄_w_* is the average connectivity strength of a given ICA component and *Z̄_b_* is the average connectivity strength between a given ICA comment and all other ICA components. Global system segregation was computed as the mean segregation across all 55 networks. Higher segregation values reflect a stronger subdivision of the brain connectome into clearly separated networks (see Fig. 1c-f for a visualization). To ensure that the parcellation scheme estimated in UK Biobank is generalizable to the Rotterdam Study sample, we repeated the high dimensional ICA, this time using data from the Rotterdam Study participants (estimating 100 components and retrieving 50 signal components) and again performed connectivity analysis on the Rotterdam-specific parcellation scheme. Resulting system segregation scores correlated highly (r=0.90) with the scores computed on the UK Biobank-based parcellation and the respective statistical results remained comparable when using either score. For consistency, we decided to report results which were retrieved from the UK Biobank-based parcellation.

### Genetic association of system segregation in UK Biobank

GWAS on system segregation was performed in the UK Biobank dataset (N=16,635) using PLINK2 (https://www.cog-genomics.org/plink/2.0/). To account for confounding effects due to population structure, the first twenty genetic principal components were considered as covariates, together with age, sex, genotype array, and assessment centre. SNP associations are reported to be significant when reaching a genome-wide significance threshold (P < 5 × 10^-8^). We identified significant independent (r^2^ < 0.6) and lead SNPs (r^2^ < 0.1) as well as genomic risk loci including all independent signals that are physically close or overlapping in a single locus (200kb). The 1000G phase 3 reference panel was used to calculate r^2^. SNP heritability was estimated using LD score regression^25^. Precalculated LD scores from the 1000 Genomes European reference population were obtained online (https://alkesgroup.broadinstitute.org/LDSCORE/).

Functional annotation: For functional annotation, we considered all independent SNPs together with all SNPs in LD with the independent SNPs and with P-values < 0.05 (N=659). We performed ANNOVAR^65^ gene-based annotation as implemented in FUMA^26^ (v.1.3.5e) using refSeq genes. In addition, CADD^27^ and RegulomeDB^31^ scores were identified for those SNPs by matching chromosome, position, reference and alternative alleles. Chromatin modification of each SNP was characterized by FUMA ChromHMM^66^. In this process, a multivariate Hidden Markov Model is used to predict 15-core chromatin states base on 5 histone marks from 127 different tissue/cell types.

Gene-based analysis: We used two strategies to identify genes related to system segregation. For positional mapping, we mapped all SNPs in the risk loci to genes based on physical distance (within a 10 Kb window) from known protein-coding genes in the human reference assembly (GRCh37/hg19). For genome-wide gene-based association analysis (GWGAS), SNP-based P-values from the GWAS output were tested with relation to the 19,427 protein-coding genes from the NCBI 37.3 gene definitions using MAGMA^33^ (v1.07). After functional annotation, 18,335 genes were covered by at least one SNP. Gene association tests were performed taking LD between SNPs into account. To account for multiple testing, we applied a stringent Bonferroni correction setting the significance threshold to P < 2.73 × 10^-6^ (i.e. 0.05/number of genes). We used all unique genes found via positional mapping and GWGAS as input for post-GWAS analyses.

Differential tissue expression: Gene expression analysis was carried out in FUMA. Input genes were tested against the GTEx.v8 data set of 54 tissue types using the hypergeometric test. Significant enrichment is reported at Bonferroni-corrected P < 9.26 × 10^-4^ (i.e. 0.05/number of tissues) and differentiating downregulated and upregulated genes dependent on the sign of the hypergeometric test.

Pathway enrichment: We used FUMA to test for relationships between genes associated with system segregation and 13,139 predefined gene sets related to biological/molecular pathways (canonical pathways: N=2,868 and GO pathways: N=10,271) which were obtained from the Molecular Signatures Database (MSigDB, v7.0). Additionally, we tested for enrichment of gene sets which were previously related to other traits as reported in the GWAS catalogue (N = 2,195). FUMA performs hypergeometric tests to evaluate whether genes of interest are overrepresented in any of the pre-defined sets. Results are reported at Bonferroni-corrected P-value taking into account the total number of gene-sets in each category.

Genetic correlation: Using LD score regression^25^, we estimated genetic correlations between system segregation and cardiovascular risk scores. SNP summary statistics for system segregation were derived from the current GWAS, while publicly available summary statistics were used for all other traits (https://nealelab.github.io/UK BiobankB_ldsc/downloads.html).

### Polygenic prediction in the Rotterdam Study

For out-of-sample prediction, we computed a PRS of system segregation for each Rotterdam Study participant using PRSice2^32^ (v2.3.3). PRS was created as a summation of an individual’s genotype data weighted by each SNP’s effect size that was estimated from the GWAS summary statistics in the UK Biobank. Clumping was carried out to further remove SNPs that are in LD with each other at r^2^ < 0.25 within a 200 bp window. P-thresholding (lowest to largest P-value: 5 × 10^-8^ − 0.5 in intervals of 5 × 10^-5^) was used to obtain a PRS with the highest possible model fit.

### Assessment of cardiovascular health

Cardiovascular risk factors were assessed for each UK Biobank participant based on Life’s Simple seven (LS7) which include blood pressure, blood cholesterol, glycaemic control, smoking status, body mass index, physical activity, and diet (Supplementary Table 7a). Following the American Heart Association’s recommendations^24^, each measure was divided into three levels (coded as poor=0, intermediate=1, and optimal=2) as detailed in Supplementary Table 7b. All scores expect of cholesterol and glycaemic control (blood samples were collected only at baseline) were available for both the baseline and the brain imaging visit. Missing raw values were imputed by multiple imputations using chained equations with 20 imputations and all remaining variables as predictors (R mice package, v3.11.0). The amount of missing data and the quality of imputation are visualized in Supplementary Fig. 6.

### Mendelian randomization

Two-sample MR analyses were carried out to assess the relationship between (1) systolic blood pressure (as exposure) and system segregation (as outcome) and (2) system segregation (as exposure) and cognition (as outcome). Genetic instruments of system segregation were derived from the current GWAS. Two additional GWAS of blood pressure and cognitive performance were conducted for the purpose of two-sample MR analysis (Supplementary Methods 1) in the largest non-overlapping samples of UK Biobank participants that had not undergone fMRI assessment and hence were not included in the current GWAS on system segregation. Genetic instruments were defined as all SNPs with P < 5 × 10^-8^ and r^2^ < 0.001 based on the European 1000 Genomes panel. As the primary method of analysis, individual MR estimates were pooled using a random-effects inverse-variance weighted (IVW) estimator^67^. MR outcome derived from the IVW method might be biased if the genetic variants are pleiotropic. As a measure of overall pleiotropy, heterogeneity in the IVW MR analyses was assessed with the Cochran’s Q statistic^68^. While it is assumed that balanced horizontal pleiotropy leads to random effects with zero means and hence no bias, directional (unbalanced) pleiotropy can lead to biased IVW estimates. To ensure that this was not the case, we computed the MR Egger intercept^68^, which should not be significantly different from zero in case of balanced pleiotropy. We additionally inspected funnel plots, which should show a symmetrical distribution. Alternative MR methods were applied, which are more robust to pleiotropy. These were the weighted median estimator^69^, the contamination-mixture method^70^, and MR-PRESSO^71^. Details about these approaches and their underlying assumptions are provided in Supplementary Methods 2. All analyses were performed in R using the MendelianRandomization (v0.4.3), TwoSampleMR (v0.5.5) and the MRPRESSO (v1.0) packages.

### Statistical analysis

All statistical analyses were performed in R v3.6.3 and a P-value below 0.05 (two-sided) was considered significant when not stated differently. All continuous values had been scaled and centred before entered into the statistical models and hence beta values represent standardized regression coefficients. We included age (data field: 21022), age^2^, sex (data field: 31), education (data field: 6138; categorized as higher (college/university degree or other professional qualification) or lower), assessment centre visited (data field: 54; applies only to UK Biobank – all Rotterdam Study data was acquired at the same centre), total GM volume (data field: 25006), and WMH volume (data field: 25781) as covariates into all statistical models unless differently stated.

### System segregation-vascular risk relationship

Multiple linear regression analyses were used to estimate the relationship between system segregation and LS7 vascular risk factors assessed at the baseline or brain imaging visit. We followed the recommended strategy for multiple imputation analysis^72^ and estimated first the statistical model in each imputed dataset (N=20) and then pooled the parameter estimates of interest into one final estimate. This strategy ensures valid results, accounting for the missing data and having the correct type I error rate. Performing the analyses in the non-imputed dataset did not change our conclusions and thus we decided to report results for the imputed dataset (N = 16,635). All models included the above-mentioned covariates as well as head motion during fMRI assessment quantified by the mean framewise displacement (data field: 25741). Since multiple tests had been performed, we Bonferroni-adjusted the P-value threshold by the number of cardiovascular risk factors, i.e. 0.05/7 = 0.007. The scores are classified on 3-levels (poor, intermediate, optimal). In order to ensure that this classification scheme has not biased our results, we additionally performed regression analyses on the continuous values. Finally, the discovered associations in the UK Biobank sample were replicated in the Rotterdam Study. Multiple linear regression models were used for this purpose including above mentioned covariates, head motion as well as signal-to-artefact ratio.

### Cognition-system segregation relationship

Linear multiple regression models were used to estimate the association between system segregation and cognitive performance in the UK Biobank (N = 7,342) and Rotterdam Study (N = 2,012). We specified a main effect model and a system segregation × age interaction model on cognitive performance. We additionally investigated the effect of system segregation on cognitive decline in a subsample of UK Biobank participants who had a follow-up cognitive assessment available (N = 2,113). To this end, we used a linear mixed-effects model to estimate a system segregation × time interaction effect on cognitive performance, while including a random intercept for each participant and the above-mentioned covariates as fixed effects.

### Two-dimensional density plots

Results of multiple linear regression analyses are displayed by two-dimensional density plots created with the geom_pointdensity function (https://github.com/LKremer/ggpointdensity). Since we used the ‘n_neighbor’ calculation with the default bandwidth, the color-code of individual points represents the number of neighbouring points.

## Data Availability

The data that supports the findings of this study were obtained from the UK Biobank under application numbers 33018 and 2532 (https://www.ukbiobank.ac.uk) and the Rotterdam Study (http://www.epib.nl/research/ergo.htm). Researchers may access these datasets upon request and in accordance with the data use agreement. PLINK (https://zzz.bwh.harvard.edu/plink/) and PRSice (https://www.prsice.info) were used for GWAS and PRS analysis. In our post-GWAS analyses, we used LDSC regression (https://github.com/bulik/ldsc), precalculated LD scores (https://alkesgroup.broadinstitute.org/LDSCORE/), FUMA (https://fuma.ctglab.nl) and MAGMA (https://ctg.cncr.nl/software/magma.

## Acknowledgements

This research has been conducted using the UK Biobank and the Rotterdam Study resources. Ethics approval for the UK Biobank study was obtained from the National Research Ethics Service Committee North West–Haydock (reference 11/NW/0382). The Rotterdam Study has been approved by the Medical Ethics Committee of the Erasmus MC (registration number MEC 02.1015) and by the Dutch Ministry of Health, Welfare and Sport (Population Screening Act WBO, license number 1071272-159521-PG. We thank all participants and all other persons who contributed to data collection, management and distribution.

The study was funded by DAAD post-doc fellowship (to JN), grants from the Alzheimer Forschung Initiative (AFI, Grant 15035 to ME), Legerlotz Stiftung (to ME), LMUexcellent (to ME); ZonMw Memorabel grant (project no 733050817 to MWV); European Union’s Horizon 2020 research and innovation programme (grant agreement no. 667375 [CoSTREAM] to MWV and MD as well as grant agreement no. 666881 [SVDs@target] to MD); Deutsche Forschungsgemeinschaft (DFG, German Research Foundation) grant for major research instrumentation (DFG, INST 409/193-1 FUGG; to MD), the DFG as part of the Munich Cluster for Systems Neurology (EXC 2145 SyNergy – ID 390857198) and the CRC 1123 (B3) to MD); Walter Benjamin-fellowship (DFG, GE 3461/1-1 to MD), Hertie Foundation for Clinical Neurosciences (to NF); LMU Förderung Forschung Lehre (Reg. 1032 to NF).

The Rotterdam Study is funded by Erasmus Medical Center and Erasmus University, Rotterdam, Netherlands Organization for the Health Research and Development (ZonMw), the Research Institute for Diseases in the Elderly (RIDE), the Ministry of Education, Culture and Science, the Ministry for Health, Welfare and Sports, the European Commission (DG XII), and the Municipality of Rotterdam. None of the funders had any role in the design and conduct of the study; collection, management, analysis, and interpretation of the data; and preparation, review, or approval of the manuscript.

