## Supplementary_Information for "Genetic variants link lower segregation of brain networks to higher blood pressure and worse cognition within the general aging population"

**Supplementary Methods 1 GWAS of blood pressure and cognition in the UK Biobank**

For the purpose of two-sample Mendelian Randomization (MR) with non-overlapping sample, we selected the largest possible UK Biobank datasets with either systolic blood pressure (N=364,061) or cognition (N = 10,558) available excluding participants with fMRI and who were thus part of the GWAS on system segregation. Systolic blood pressure was adjusted for anti-hypertensive medication[1]. Two separate GWAS were run for blood pressure and cognitive performance using PLINK2 (https://www.cog-genomics.org/plink/2.0/) and accounting for the first twenty genetic principle components, together with age, sex, genotype array and assessment center. Based on the GWAS output of blood pressure, we selected SNPs at $P < 5 \times 10^{-8}$ as instruments and clumped for linkage disequilibrium (LD) to $r^2 < 0.001$ based on the European 1000 Genomes panel. These variants (N = 312) were then used as the genetic instruments of blood pressure in the two-sample MR analysis. From the GWAS output of cognitive performance, we extracted beta-values and standard errors for the 10 genetic instruments of system segregation identified in the current GWAS which were then entered into two-sample MR analysis.

**Supplementary Methods 2 MR analysis methods**

The weighted median estimator allows the use of invalid instruments as long as at least half of the instruments used in the MR analysis are valid[2]. The contamination mixture method constructs a likelihood function of the individual estimates and under the assumption that the estimates of the valid instruments would follow a distribution centered around the causal effect and any invalid instruments would follow a distribution around zero, it calculates MR estimates that would maximize this likelihood[3]. The contamination method assumes that only some of the genetic variants used are valid instruments and it has been found to perform better than other methods under the presence of invalid instruments[4]. Finally, MR-PRESSO regresses the SNP-outcome estimates against the SNP-exposure estimates to test for outlier SNPs[5]. Outliers are detected by sequentially removing all variants from the analyses and comparing the residual sum of squares as a global measure of heterogeneity ($p<0.05$ for detecting outliers); outliers are then removed and outlier-corrected estimates are provided[5]. Although outlier-robust, MR-PRESSO still relies on the assumption that at least half of the variants are valid instruments[5].

**Supplementary Table 1 Characteristics of subsamples**

| Cohort | UK Biobank | | | | Rotterdam Study | |
|---|---|---|---|---|---|---|
| **Analysis** | GWAS system segregation, MR | GWAS cognition MR analysis | GWAS blood pressure, MR analysis | System segregation - cognition association | Polygenic prediction system segregation | System segregation - cognition association |
| **Inclusion criteria** | Genetics fMRI | Genetics cognition no fMRI | Genetics blood pressure no fMRI | Cognition fMRI | Genetics fMRI | Cognition fMRI |
| **N** | 16,635 | 10,558 | 364,061 | 7,342 | 2,414 | 2,012 |
| **Age, y** | 63.14(7.44) | 64.79(7.37) | 57.69(7.93) | 63.06(7.28) | 67.22(9.04) | 65.72(8.68) |
| **Sex, women** | 8,799(52.89) | 5,271(49.92) | 194,757(53.45) | 3,858(52.55) | 1,292(53.52) | 1,078(53.58) |
| **Education, college degree** | 7,186(43.20) | 4,944(46.83) | 110,572(30.34) | 3,462(47.15) | 655(27.14) | 590(29.32) |
| **(Symbol/Letter)-Digit Substitution** | Na | 18.87(5.18) | Na | 19.51(5.14) | Na | 30.92(6.58) |
| **Total GM volume, mL** | 617.66(55.22) | Na | Na | 621.92(55.48) | 534.81(62.07) | 535.53(59.93) |
| **Total WMH, mL** | 4.55(5.86) | Na | Na | 4.92(5.89) | 5.87(9.19) | 5.49(8.72) |
| **Head motion fMRI, FD** | 0.12(0.06) | Na | Na | 0.12(0.06) | 0.07(0.03) | 0.07(0.03) |
| **Systolic BP, mmHg** | 138.83(18.94) | 142.42(19.83) | 141.01(19.66) | 140(19.07) | 136.63(19.10) | 136.06(19.02) |
| **Diastolic BP, mmHg** | 78.54(10.55) | 79.01(10.59) | 82.54(10.67) | 78.39(10.56) | 79.50(11.21) | 80.40(10.98) |
| **Body mass index, kg/m$^2$** | 26.58(4.39) | 26.62(4.56) | 27.49(4.78) | 26.43(4.34) | 27.13(3.91) | 27.15(3.95) |
| **Smoking, never** | 10,460(62.88) | 6,648(62.97) | 196,857(54.02) | 4,712(64.18) | 813(33.68) | 743(36.92) |
| **Smoking, previous** | 5,579(33.54) | 3.526(33.40) | 129,197(35.46) | 2,366(32.23) | 1,398(57.90) | 1101(54.74) |
| **Smoking, current** | 596(3.58) | 332(3.14) | 37,025(10.16) | 223(3.04) | 203(8.42) | 168(8.34) |

BP blood pressure, FD frame-wise displacement, GM grey matter, y years, NA not applicable, WMH white matter hyperintensities

**Supplementary Table 2 Nine independent genomic loci associated with system segregation**

| Locus | Chr | Position | P-value | Start | End | Independent sig. SNPs | Lead SNPs | Mapped Genes |
|---|---|---|---|---|---|---|---|---|
| **1** | 6 | 1366718 | 1.57e-09 | 1364471 | 1381066 | rs7766042 | rs7766042 | FOXF2 |
| **2** | 6 | 96894135 | 4.89e-16 | 95894255 | 97895889 | rs13208422;rs11152951;rs10457146;rs9386670;rs2971606;rs2025044 | rs11152951 | UFL1;FHL5; GPR63;NDUF;AF4;KLHL32; MMS22L |
| **3** | 9 | 71426328 | 1.09e-12 | 71370174 | 71515259 | rs11143801;rs7875396 | rs11143801 | PIP5K1B; FAM122A |
| **4** | 10 | 96039597 | 1.32e-28 | 95196591 | 97108669 | rs536348722;rs2274224;rs11187844;rs11187850;rs3758526;rs11187830;rs60341058;rs3740360;rs7910752;rs75017201;rs564376090; rs7092226 | rs2274224 | PLCE1;CEP55;NOC3L;TBC1D12;HELLS;CYP2C18;CYP2C19;CYP2C9;C10orf129;SORBS1 |
| **5** | 10 | 134282867 | 1.00e-33 | 134269313 | 134517013 | rs138004790;rs9645539;rs7907962;rs4486544;rs4880389;rs4880397;rs7076422;rs11146402;rs28454681;rs10747058;rs10781575;rs1133400;rs11146456 | rs138004790; rs9645539; rs1133400 | INPP5A;C10orf91 |
| **6** | 11 | 10676255 | 3.85e-11 | 9759616 | 11705923 | rs1544863;rs11042933;rs11042940;rs730322;rs10840469 | rs1544863 | SBF2;AMPD3; RNF141;LYVE1;MRVI1;CTR9;EIF4G2; ZBED5;GALNT18 |
| **7** | 11 | 70040645 | 4.89e-12 | 69058825 | 70981648 | rs4980704;rs34094842;rs3781658;rs7127129;rs76136259;rs10898840 | rs4980704;rs34094842 | CCND1;ORAOV1;ANO1;FADD;SHANK2 |
| **8** | 11 | 101710122 | 6.43e-11 | 100722966 | 102698257 | rs1954765;rs2155059;rs12282371;rs10791530;rs11224992;rs10895201; rs732257 | rs12282371 | TMEM133;PGR;TRPC6;ANGPTL5;KIAA1377;YAP1;BIRC3;BIRC2;TMEM123; RP11-315O6.2; MMP7;MMP20;MMP27;MMP1;MMP3 |
| **9** | 17 | 19138174 | 5.68e-11 | 18965491 | 19299144 | rs72639204 | rs72639204 | EPN2;B9D1;MAPK7;MFAP4 |

**Supplementary Table 3 Characteristics of the 12 lead SNPs associated with system segregation**

| No | Locus | uniqID | rsID | p | beta | se | MAF | Func. position | Chromatin | RDB | CADD |
|---|---|---|---|---|---|---|---|---|---|---|---|
| 1 | 1 | 6:1366718:C:T | rs7766042 | 1,57E-09 | -0,05868 | 0,009714 | 0,07753 | intergenic | 2 | 6 | 1,899 |
| 2 | 2 | 6:96894135:C:G | rs11152951 | 4,89E-16 | -0,06293 | 0,007748 | 0,2087 | ncRNA_intronic | 9 | 7 | 4,978 |
| 3 | 3 | 9:71426328:C:T | rs11143801 | 1,09E-12 | 0,043907 | 0,006163 | 0,3588 | intronic | 4 | 7 | 2,617 |
| 4 | 4 | 10:96039597:C:G | rs2274224 | 1,32E-28 | 0,066674 | 0,005998 | 0,4563 | exonic | 4 | 6 | 17,35 |
| 5 | 5 | 10:134270762:G:T | rs138004790 | 4,07E-08 | -0,06968 | 0,012692 | 0,07753 | intergenic | 4 | 2b | 1,501 |
| 6 | 5 | 10:134282867:G:T | rs9645539 | 1E-33 | -0,07265 | 0,005989 | 0,4781 | intergenic | 2 | 3a | 1,603 |
| 7 | 5 | 10:134459388:A:G | rs1133400 | 5,97E-12 | -0,04935 | 0,007168 | 0,2137 | exonic | 4 | 4 | 17,57 |
| 8 | 6 | 11:10676255:C:T | rs1544863 | 3,85E-11 | 0,045111 | 0,00682 | 0,2614 | intronic | 4 | 5 | 0,359 |
| 9 | 7 | 11:69823461:A:G | rs4980704 | 2,37E-08 | 0,033443 | 0,005987 | 0,4493 | intergenic | 5 | 5 | 1,926 |
| 10 | 7 | 11:69983047:A:G | rs34094842 | 1,88E-09 | 0,040266 | 0,006699 | 0,334 | ncRNA_intronic | 4 | 5 | 2,112 |
| 11 | 8 | 11:101710122:A:G | rs12282371 | 6,43E-11 | 0,048437 | 0,007409 | 0,1968 | NA | 14 | 7 | 0,301 |
| 12 | 9 | 17:19138174:C:T | rs72639204 | 5,68E-11 | -0,05093 | 0,007768 | 0,1928 | intronic | 5 | 5 | 4,188 |

**Supplementary Table 4 SNPs included in the best-fit polygenic risk score of system segregation in the Rotterdam Study sample**

| No | uniqID | rsID | p | beta | se |
|---|---|---|---|---|---|
| 1 | 6:1366718:C:T | rs7766042 | 1.57e-09 | -0.059 | 0.020 |
| 2 | 6:96904366:C:T | rs4078038 | 6.11e-16 | -0.063 | 0.008 |
| 3 | 9:71426328:C:T | rs11143801 | 1.09e-12 | 0.044 | 0.006 |
| 4 | 10:96023077:C:T | rs57866767 | 2.01e-28 | 0.066 | 0.006 |
| 5 | 10:96104665:G:T | rs3758526 | 1.26e-10 | -0.058 | 0.009 |
| 6 | 10:134270762:G:T | rs138004790 | 4.07e-08 | -0.070 | 0.013 |
| 7 | 10:134282867:G:T | rs9645539 | 1.00e-33 | -0.073 | 0.006 |
| 8 | 10:134459388:A:G | rs1133400 | 5.97e-12 | -0.049 | 0.007 |
| 9 | 11:10676255:C:T | rs1544863 | 3.85e-11 | 0.045 | 0.007 |
| 10 | 11:69823461:A:G | rs4980704 | 2.37e-08 | 0.033 | 0.006 |
| 11 | 11:69983047:A:G | rs34094842 | 1.88e-09 | 0.040 | 0.007 |
| 12 | 11:101710122:A:G | rs12282371 | 6.43e-11 | 0.048 | 0.007 |
| 13 | 17:19138174:C:T | rs72639204 | 5.68e-11 | -0.051 | 0.008 |

**Supplementary Table 5a Results of pathway enrichment analysis for canonical pathways (N = 2,868)**

| GeneSet | N | n | P-value | adjusted P | genes |
|---|---|---|---|---|---|
| REACTOME_COLLAGEN_DEGRADATION | 63 | 4 | 6,22e-06 | 0,006187 | MMP7:MMP20:MMP1:MMP3 |
| REACTOME_REGULATED_NECROSIS | 20 | 3 | 7,05e-06 | 0,006187 | FADD:BIRC3:BIRC2 |
| PID_P75_NTR_PATHWAY | 68 | 4 | 8,44e-06 | 0,006187 | BIRC3:BIRC2:MMP7:MMP3 |
| REACTOME_XENOBIOTICS | 24 | 3 | 1,25e-05 | 0,006845 | CYP2C18:CYP2C19:CYP2C9 |
| REACTOME_COLLAGEN_DEGRADATION | 63 | 4 | 6,22e-06 | 0,006187 | MMP7:MMP20:MMP1:MMP3 |

**Supplementary Table 5b Results of pathway enrichment analysis for GO pathways (N = 10,271)**

| GeneSet | N | n | P-value | adjusted P | genes |
|---|---|---|---|---|---|
| GO_COLLAGEN_CATABOLIC_PROCESS | 46 | 5 | 2.59e-08 | 0,00019 | MMP7:MMP20:MMP27:MMP1:MMP3 |
| GO_METALLOENDOPEPTIDASE_ACTIVITY | 101 | 5 | 1.38e-06 | 0,00227 | MMP7:MMP20:MMP27:MMP1:MMP3 |
| GO_RESPONSE_TO_DRUG | 1008 | 11 | 2.60e-06 | 0,008195 | CYP2C18:CYP2C19:CYP2C9:CCND1:FADD:TRPC6:YAP1:BIRC2:MMP3:[illegible] |
| GO_ARACHIDONIC_ACID_MONOOXYGENASE_ACTIVITY | 16 | 3 | 3.48e-06 | 0,002864 | CYP2C18:CYP2C19:CYP2C9 |
| GO_NEG._REGULATION_NECROTIC_CELL_DEATH | 17 | 3 | 4.22e-06 | 0,008195 | FADD:BIRC3:BIRC2 |
| GO_NECROTIC_CELL_DEATH | 58 | 4 | 4.46e-06 | 0,008195 | FADD:BIRC3:BIRC2:TMEM123 |
| GO_COLLAGEN_CATABOLIC_PROCESS | 46 | 5 | 2.59e-08 | 0,00019 | MMP7:MMP20:MMP27:MMP1:MMP3 |

**Supplementary Table 6 Results of pathway enrichment analysis for GWAS catalogue reported genes (N = 2,195)**

| GeneSet | N | n | P-value | adjusted P | genes |
|---|---|---|---|---|---|
| Plasma clozapine-norclozapine ratio in treatment-resistant schizophrenia | 16 | 7 | 1.80e-14 | 3.28e-11 | PLCE1, NOC3L, TBC1D12, HELLS, CYP2C18, CYP2C19, CYP2C9 |
| Migraine | 76 | 8 | 6.91e-11 | 6.27e-08 | PLCE1, INPP5A, MRVI1, CTR9, YAP1, FUT9, UFL1, FHL5 |
| Diastolic blood pressure | 592 | 13 | 1.83e-08 | 1.11e-05 | PLCE1, CYP2C19, CYP2C9, SWAP70, AMPD3, MYEOV, CCND1, ANO1, ARHGAP42, TMEM133, PGR, EPN2, FUT9, UFL1 |
| Thiopurine-induced alopecia in inflammatory bowel disease | 11 | 4 | 2.46e-08 | 1.12e-05 | TBC1D12, HELLS, CYP2C18, CYP2C19 |
| Pulse pressure | 647 | 12 | 4.31e-07 | 1.57e-04 | PLCE1, CYP2C19, CYP2C9, INPP5A, SWAP70, AMPD3, MYEOV, ARHGAP42, TMEM133, YAP1, FUT9, UFL1, FHL5 |
| Clopidogrel active metabolite levels | 7 | 3 | 9.16e-07 | 2.77e-04 | CYP2C18, CYP2C19, CYP2C9 |
| Warfarin maintenance dose | 10 | 3 | 3.12e-06 | 7.67e-04 | CYP2C18, CYP2C19, CYP2C9 |
| Prostate cancer | 302 | 8 | 3.38e-06 | 7.67e-04 | MYEOV, YAP1, BIRC3, BIRC2, TMEM123, MMP7, MMP20, MMP27 |
| Systolic blood pressure | 738 | 11 | 1.13e-05 | 2.22e-03 | PLCE1, CYP2C19, CYP2C9, INPP5A, SWAP70, AMPD3, ANO1, ARHGAP42, TMEM133, FUT9, UFL1, PIP5K1B |

**Supplementary Table 7a UK Biobank data fields used in the construction of the Life's Simple 7 cardiovascular risk score**

| Life's simple 7 item | Data fields used |
|---|---|
| Smoking status | 20116 |
| BMI | 21001 |
| Physical activity | 884, 904 |
| Diet | 1309, 1319, 1289, 1299, 1448, 1438, 1468, 1458, 1329, 1339, 1408, 1418, 1428, 2654, 1349, 1359, 1369, 1379,1389, 3680 |
| Blood pressure | 4080, 4079 |
| Cholesterol levels | 30780 |
| Glycemic control | 30750 |

**Supplementary Table 7b. Definition of Life's Simple 7 sub-scores in the UK Biobank**

| | Blood Pressure | Cholesterol levels** | Glycemic control | Smoking status | BMI | Physical activity | Diet* |
|---|---|---|---|---|---|---|---|
| **Optimal (score=2)** | SBP <120 mm Hg and DBP <80 mm Hg untreated | LDL-C <130 mg/dl | HbA1c <5.7% | Never smoked | <25 kg/m$^2$ | >4 days/week of moderate/vigorous physical activity | >7 |
| **Intermediate (score=1)** | SBP 120-139 or DBP 80-89 mm Hg OR SBP <120 mm Hg and DBP <80 mm Hg treated | LDL-C 130-159 mg/dl OR LDL-C <130 mg/dl treated | HbA1c 5.7-6.4% OR HbA1c <5.7% treated | Former smoker | 25-29.9 kg/m$^2$ | ≤4 days/week of moderate/vigorous physical activity | 4-7 |
| **Poor (score=0)** | SBP ≥140 mm Hg or DBP ≥90 mm Hg | LDL-C ≥ 160 mg/dl | HbA1c ≥ 6.4% | Current smoker | ≥30 kg/m$^2$ | No moderate/ vigorous physical activity | <4 |

* Healthy diet score according to Mozaffarian[6] and Said et al.[7]; higher scores indicate adherence to a healthier diet for prevention of cardiovascular disease.

BMI body mass index, DBP diastolic blood pressure, HbA1c glycated hemoglobin, LDL-C low-density lipoprotein cholesterol, SBP systolic blood pressure,

**Supplementary Table 8. Genetic correlation between system segregation and LS7 cardiovascular risk factors using LD score regression**

| LS7 cardiovascular health traits | UKB ID | rg | se | z | p | h2_obs | h2_obs_se | h2_int | h2_int_se | gcov_int | gcov_int_se |
|---|---|---|---|---|---|---|---|---|---|---|---|
| **Systolic blood pressure** | **4080** | **-0,1783** | **0,0545** | **-3,2718** | **0,0011** | 0,1436 | 0,006 | 1,0773 | 0,0181 | -0,005 | 0,0067 |
| Diastolic blood pressure | 4079 | -0,1453 | 0,0594 | -2,4462 | **0,0144** | 0,1331 | 0,0054 | 1,071 | 0,0184 | -0,0011 | 0,0065 |
| LDL direct | 30780 | 0,0025 | 0,0979 | 0,0251 | 0,98 | 0,0464 | 0,0489 | 1,4085 | 0,3338 | 0,0011 | 0,0073 |
| Glycated haemoglobin (HbA1c) | 30750 | -0,0104 | 0,049 | -0,2119 | 0,8322 | 0,1808 | 0,016 | 1,2585 | 0,1007 | -0,0015 | 0,0073 |
| Smoking, never | 20116 | -0,0575 | 0,0508 | -1,1307 | 0,2582 | 0,0952 | 0,0041 | 1,041 | 0,0114 | 0,0019 | 0,0058 |
| Body mass index | 21001 | -0,0026 | 0,05 | -0,0525 | 0,9581 | 0,2475 | 0,0094 | 1,0655 | 0,0199 | 0,0128 | 0,0066 |
| Number of days/week of moderate physical activity | 884 | 0,0658 | 0,0699 | 0,9411 | 0,3467 | 0,0404 | 0,0025 | 1,005 | 0,0089 | -0,0049 | 0,0051 |
| Number of days/week of vigorous physical activity | 904 | 0,1525 | 0,0713 | 2,1376 | **0,0325** | 0,0362 | 0,0026 | 1,0174 | 0,0097 | -0,0145 | 0,0054 |
| Cooked vegetable intake | 1289 | 0,0185 | 0,0732 | 0,2524 | 0,8008 | 0,0399 | 0,0021 | 1,0125 | 0,0078 | 0,0031 | 0,0055 |
| Salad / Raw vegetable intake | 1299 | -0,0348 | 0,0703 | -0,4945 | 0,621 | 0,0434 | 0,0026 | 1,0092 | 0,0091 | 0,0005 | 0,0053 |
| Fresh fruit intake | 1309 | 0,0132 | 0,06 | 0,2193 | 0,8264 | 0,0614 | 0,0035 | 1,005 | 0,0111 | -0,0082 | 0,0061 |
| Dried fruit intake | 1319 | 0,0947 | 0,0559 | 1,6947 | 0,0901 | 0,0637 | 0,003 | 1,0171 | 0,0091 | -0,015 | 0,0055 |
| Oily fish intake | 1329 | -0,0051 | 0,0611 | -0,0828 | 0,934 | 0,0643 | 0,0035 | 1,0048 | 0,0113 | -0,0121 | 0,0056 |
| Non-oily fish intake | 1339 | 0,0041 | 0,0882 | 0,0465 | 0,9629 | 0,0256 | 0,002 | 1,0131 | 0,0089 | -0,007 | 0,0056 |
| Processed meat intake | 1349 | -0,0872 | 0,0653 | -1,3347 | 0,182 | 0,0437 | 0,0031 | 1,0168 | 0,0104 | 0,0052 | 0,0051 |
| Poultry intake | 1359 | 0,0138 | 0,0732 | 0,1882 | 0,8507 | 0,0318 | 0,0022 | 1,0084 | 0,009 | -0,0048 | 0,0053 |
| Beef intake | 1369 | -0,0252 | 0,0712 | -0,354 | 0,7233 | 0,037 | 0,0026 | 1,0282 | 0,0098 | 0,0023 | 0,0056 |
| Lamb/mutton intake | 1379 | -0,1156 | 0,0657 | -1,7599 | 0,0784 | 0,042 | 0,0025 | 1,0254 | 0,0092 | 0,0115 | 0,0053 |
| Pork intake | 1389 | 0,0167 | 0,0713 | 0,2349 | 0,8143 | 0,0296 | 0,0022 | 1,0157 | 0,0081 | -0,0034 | 0,0054 |
| Cheese intake | 1408 | -0,008 | 0,0704 | -0,1137 | 0,9094 | 0,0681 | 0,0034 | 1,0186 | 0,0111 | 0,0064 | 0,0056 |
| Milk type: semi-skimmed | 1418 | 0,0824 | 0,0991 | 0,8312 | 0,4059 | 0,0138 | 0,0018 | 1,0066 | 0,0077 | -0,0004 | 0,0053 |
| Bread intake | 1438 | -0,0352 | 0,0695 | -0,5059 | 0,6129 | 0,0496 | 0,0028 | 1,0069 | 0,009 | 0,0038 | 0,0057 |
| Bread type: wholegrain | 1448 | 0,1019 | 0,0664 | 1,5337 | 0,1251 | 0,0472 | 0,0025 | 1,033 | 0,009 | -0,0036 | 0,0057 |
| Cereal intake | 1458 | -0,0038 | 0,0644 | -0,0584 | 0,9534 | 0,0597 | 0,0028 | 1,018 | 0,01 | -0,0063 | 0,006 |
| Cereal type: oat | 1468 | 0,0998 | 0,0978 | 1,0211 | 0,3072 | 0,0186 | 0,0021 | 1,0099 | 0,0078 | -0,008 | 0,0048 |
| Spread type: butter | 1428 | 0,053 | 0,0649 | 0,8163 | 0,4143 | 0,0435 | 0,0022 | 0,9879 | 0,0093 | 0,0009 | 0,0052 |
| Non-butter spread type: margarine | 2654 | 0,1019 | 0,1003 | 1,0159 | 0,3097 | 0,0266 | 0,003 | 1,0085 | 0,0077 | -0,0074 | 0,0051 |

**Supplementary Table 9a Observational association between system segregation and cardiovascular risk factors at baseline (N = 16,635)**

| LS7 sub-score | Contrast | Estimate | Std error | T-value | DF | P-value |
|---|---|---|---|---|---|---|
| Blood pressure | poor vs. intermediate | 0.093 | 0.014 | 6.544 | 6814.137 | <0.001 |
| | poor vs. optimal | 0.141 | 0.020 | 7.212 | 9168.980 | <0.001 |
| Cholesterol | poor vs. intermediate | 0.016 | 0.016 | 0.995 | 5770.282 | 0.320 |
| | poor vs. optimal | 0.038 | 0.017 | 2.167 | 4294.087 | 0.030 |
| Glycaemic control | poor vs. intermediate | -0.068 | 0.040 | -1.712 | 9576.839 | 0.087 |
| | poor vs. optimal | -0.033 | 0.035 | -0.946 | 12195.956 | 0.344 |
| Smoking | poor vs. intermediate | 0.011 | 0.028 | 0.396 | 16360.056 | 0.692 |
| | poor vs. optimal | 0.017 | 0.027 | 0.623 | 16446.509 | 0.533 |
| Body mass index | poor vs. intermediate | -0.008 | 0.019 | -0.444 | 16568.094 | 0.657 |
| | poor vs. optimal | 0.026 | 0.021 | 1.238 | 16510.278 | 0.216 |
| Physical activity | poor vs. intermediate | -0.012 | 0.021 | -0.589 | 16258.384 | 0.556 |
| | poor vs. optimal | -0.001 | 0.024 | -0.044 | 16382.976 | 0.965 |
| Diet | poor vs. intermediate | 0.007 | 0.018 | 0.398 | 309.797 | 0.691 |
| | poor vs. optimal | 0.001 | 0.031 | 0.032 | 2273.650 | 0.974 |

**Supplementary Table 9b Observational association between system segregation and cardiovascular risk factors at imaging (N = 16,635)**

| LS7 sub-score | Contrast | Estimate | Std error | T-value | DF | P-value |
|---|---|---|---|---|---|---|
| Blood pressure | poor vs. intermediate | 0.085 | 0.015 | 5.556 | 560.037 | <0.001 |
| | poor vs. optimal | 0.159 | 0.021 | 7.685 | 1232.870 | <0.001 |
| Smoking | poor vs. intermediate | 0.030 | 0.035 | 0.852 | 14771.889 | 0.394 |
| | poor vs. optimal | 0.030 | 0.034 | 0.885 | 14208.275 | 0.376 |
| Body mass index | poor vs. intermediate | -0.018 | 0.020 | -0.937 | 10689.666 | 0.349 |
| | poor vs. optimal | 0.022 | 0.022 | 0.979 | 6569.506 | 0.328 |
| Physical activity | poor vs. intermediate | 0.015 | 0.025 | 0.589 | 10916.402 | 0.556 |
| | poor vs. optimal | 0.013 | 0.027 | 0.491 | 9820.529 | 0.624 |
| Diet | poor vs. intermediate | 0.009 | 0.017 | 0.536 | 394.865 | 0.592 |
| | poor vs. optimal | 0.050 | 0.035 | 1.439 | 923.975 | 0.150 |

**Supplementary Table 10 Results of two-sample Mendelian Randomization analyses in the UK Biobank**

| Exposure – Outcome | Method | Standardized estimate | CI 95% | P |
|---|---|---|---|---|
| **Blood pressure** (non-imaging sample, N 364,061) – **System segregation** (imaging sample, N 16,638) | IVW | -0.144 | -0.228, -0.060 | 0.001 |
| | ConMix | -0.118 | -0.194, -0.026 | 0.014 |
| | MR-PRESSO[2] | -0.119 | -0.173, -0.117 | 0.002 |
| | Weighted median | -0.101 | -0.204, -0.001 | 0.052 |
| | Heterogeneity test[1] | 487.1 | | <0.001 |
| **System segregation** (imaging sample, N 16,638) – **Cognition** (non-imaging sample, N 10,558) | IVW | 0.104 | 0.028, 0.181 | 0.008 |
| | ConMix | 0.106 | 0.030, 0.181 | 0.006 |
| | MR-PRESSO[3] | 0.106 | 0.048, 0.164 | 0.006 |
| | Weighted median | 0.144 | 0.051, 0.238 | 0.002 |
| | Heterogeneity test[1] | 5.082 | | 0.827 |

[1] Cochran's Q statistic: significant results indicate that one or more instruments are likely to be pleiotropic, [2] outlier corrected estimate, [3] MR-PRESSO detected no outliers

**Supplementary Table 11a Parameter estimates and goodness of fit measures for confirmatory factor analysis in the UK Biobank**

| Latent factor | Cognitive test | B | SE | Z | P | Beta |
|---|---|---|---|---|---|---|
| General cognitive abilities "g-factor" | Fluid Intelligence | 1.00 | 0.000 | NA | NA | 0.598 |
| | Symbol Digit Substitution | 0.969 | 0.016 | 59.404 | <0.001 | 0.579 |
| | Trail Making Test, part B | 1.244 | 0.019 | 66.897 | <0.001 | 0.744 |
| | Matrix Pattern Completion | 0.985 | 0.016 | 60.077 | <0.001 | 0.589 |
| | Pairs Matching | 0.495 | 0.014 | 34.413 | <0.001 | 0.296 |
| | Numeric Memory | 0.742 | 0.015 | 48.724 | <0.001 | 0.444 |

Adjusted goodness of fit index (agfi>0.90[*])=0.960; comparative fit index (cfi>0.90)=0.952, root mean square error of approximation (rmsea<0.08)=0.074 (CI:0.070-0.078), standardized room mean square residual (srmr<0.08)=0.033

**Supplementary Table 11b Parameter estimates and goodness of fit measures for confirmatory factor analysis in the Rotterdam Study**

| Latent factor | Cognitive test | B | SE | Z | P | Beta |
|---|---|---|---|---|---|---|
| General cognitive abilities "g-factor" | Stroop Task | 1.000 | 0.000 | NA | NA | 0.727 |
| | Delayed Recall | 0.757 | 0.033 | 22.782 | <0.001 | 0.540 |
| | Letter Digit Substitution | 1.075 | 0.036 | 30.029 | <0.001 | 0.785 |
| | Verbal Fluency | 0.779 | 0.033 | 23.498 | <0.001 | 0.558 |
| | Purdue Pegboard | 0.803 | 0.033 | 24.344 | <0.001 | 0.581 |

Adjusted goodness of fit index (agfi>0.90[*])=0.979; comparative fit index (cfi>0.90)=0.987, root mean square error of approximation (rmsea<0.08)=0.056 (CI:0.02-0.072), standardized room mean square residual (srmr<0.08)=0.022

[*]cut-off for good fit is indicated in brackets based on previous recommendations[8]

### Supplementary Figures

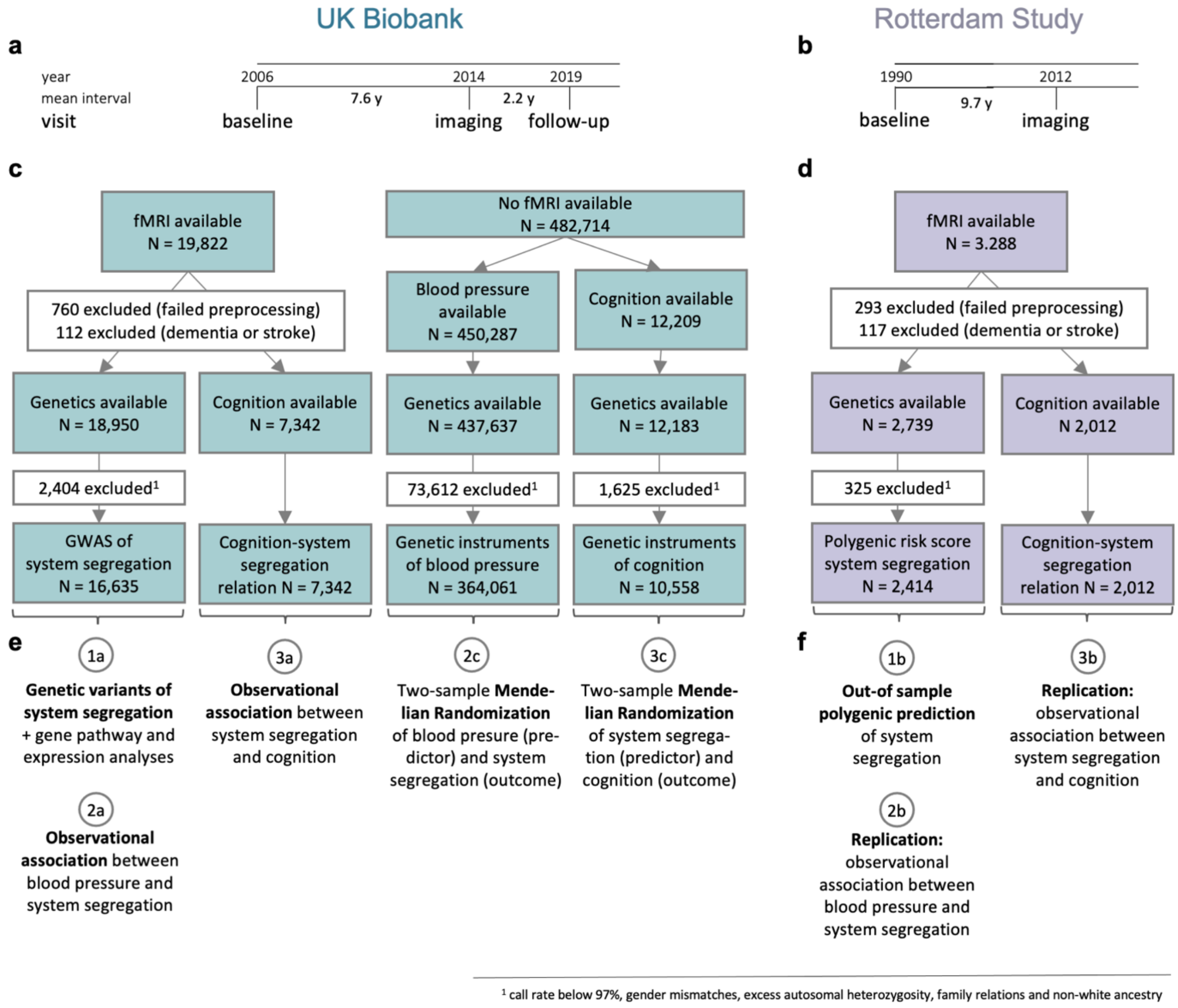


**Supplementary Fig. 1 Overview of the study design, data selection and analysis strategy**
The timelines show in which year the study relevant visits started to take place in **a** the UK Biobank and in **b** the Rotterdam Study cohort. For the UK Biobank, genetics and blood pressure had been collected at baseline. At the imaging visit, system segregation, blood pressure and cognition were assessed, while cognition was re-assessed during the follow-up visit. For the Rotterdam Study, genetic data was acquired at baseline and system segregation, blood pressure and cognition were assessed at the imaging visit. The flow diagrams illustrate the selection and exclusion of in **c** UK Biobank data (availability as of March 2019) and in **d** Rotterdam Study data (availability as of December 2019). Since we were interested in the genetic and risk factors of system segregation, the key analyses had been performed in participants with fMRI available. UK Biobank participants without fMRI available were only studied for the purpose of two-sample Mendelian randomization with non-overlapping samples. The lower panel visualizes in **e** the main analyses in the UK Biobank and in **f** their replications in the Rotterdam Study cohort.

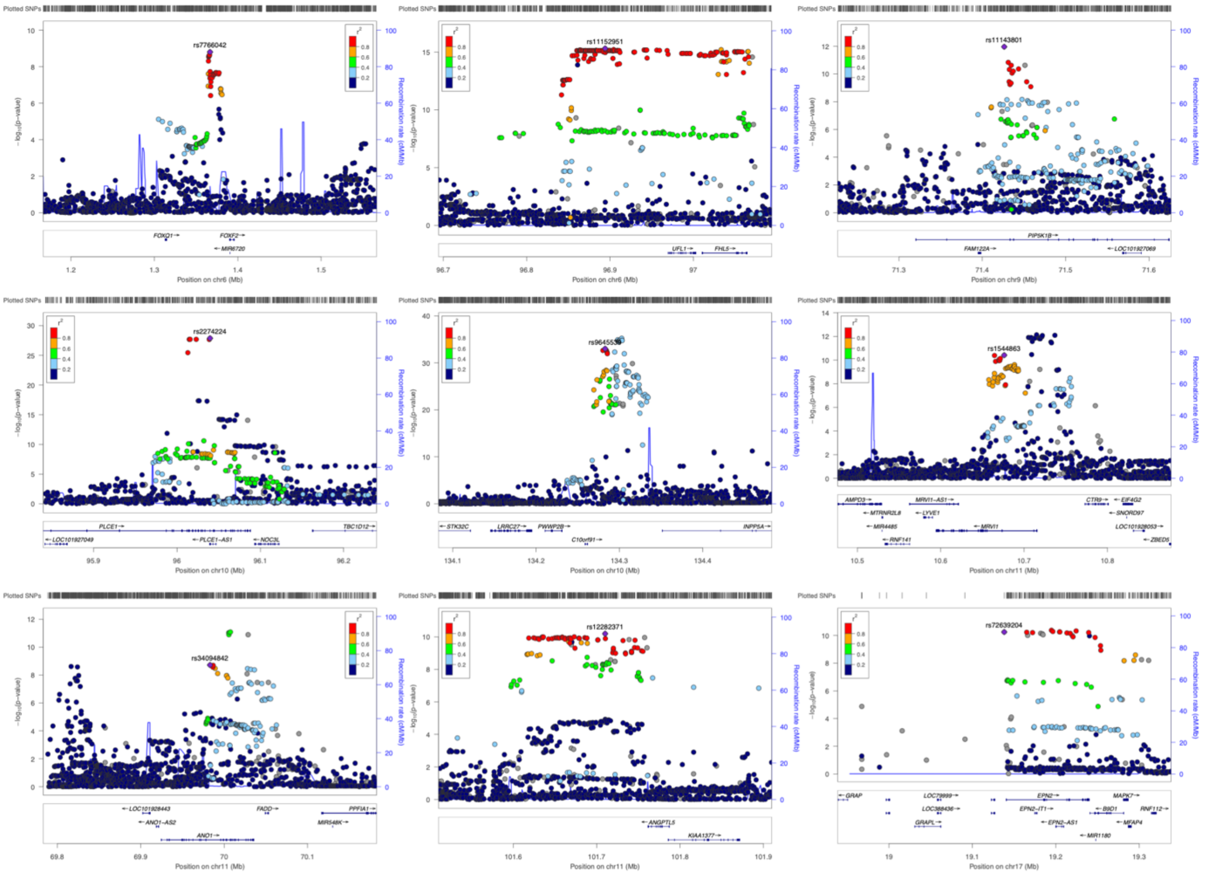


**Supplementary Fig. 2 Visualization of the nine independent risk loci associated with system segregation**

The locus zoom plots illustrate the association with system segregation (left y-axis; log10-transformed p-values) and the position of the identified genomic risk loci on the respective chromosome (x-axis). The blue line shows the recombination rate (right y-axis). For each risk locus, the top lead SNP is annotated and marked by a purple diamond, while all other SNPs are colored based on their correlation ($r^2$) with the labelled top SNP. Positions of genes are displayed below the plot. Arrows besides the gene names show the direction of transcription. Detailed information about each risk locus including lead SNPs, mapped genes, and statistical results can be found in Supplementary Table 2.

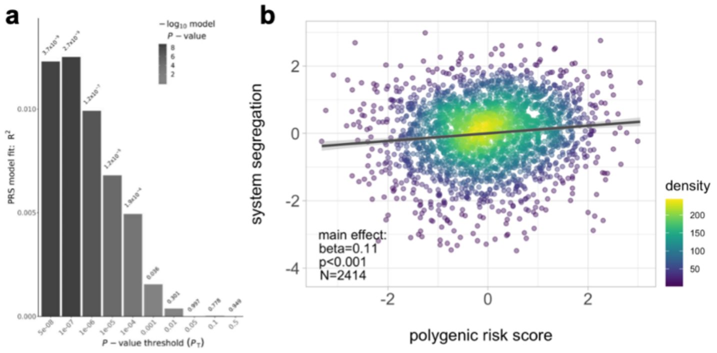


**Supplementary Fig. 3 Polygenic prediction of system segregation in the Rotterdam Study sample (N=2,414)**
**a** The bar plot shows the variance explained in system segregation (y-axis) by different polygenic risk scores (PRS) which were computed across a range of different P-value thresholds for SNP inclusion (x-axis). The SNP-based effect sizes were derived from the current GWAS results performed in the UK Biobank sample. **b** The scatter plot illustrates the main effect of the best fitted z-transformed PRS (x-axis) and z-transformed system segregation (y-axis). Statistical results were derived from multiple regression analysis including age, age$^2$, sex, education, grey matter volume, white matter hyperintensities volume, head motion during fMRI (i.e. frame-wise displacement) and signal-to-artefact ratio as covariates. Linear model fits are indicated together with 95% confidence intervals.

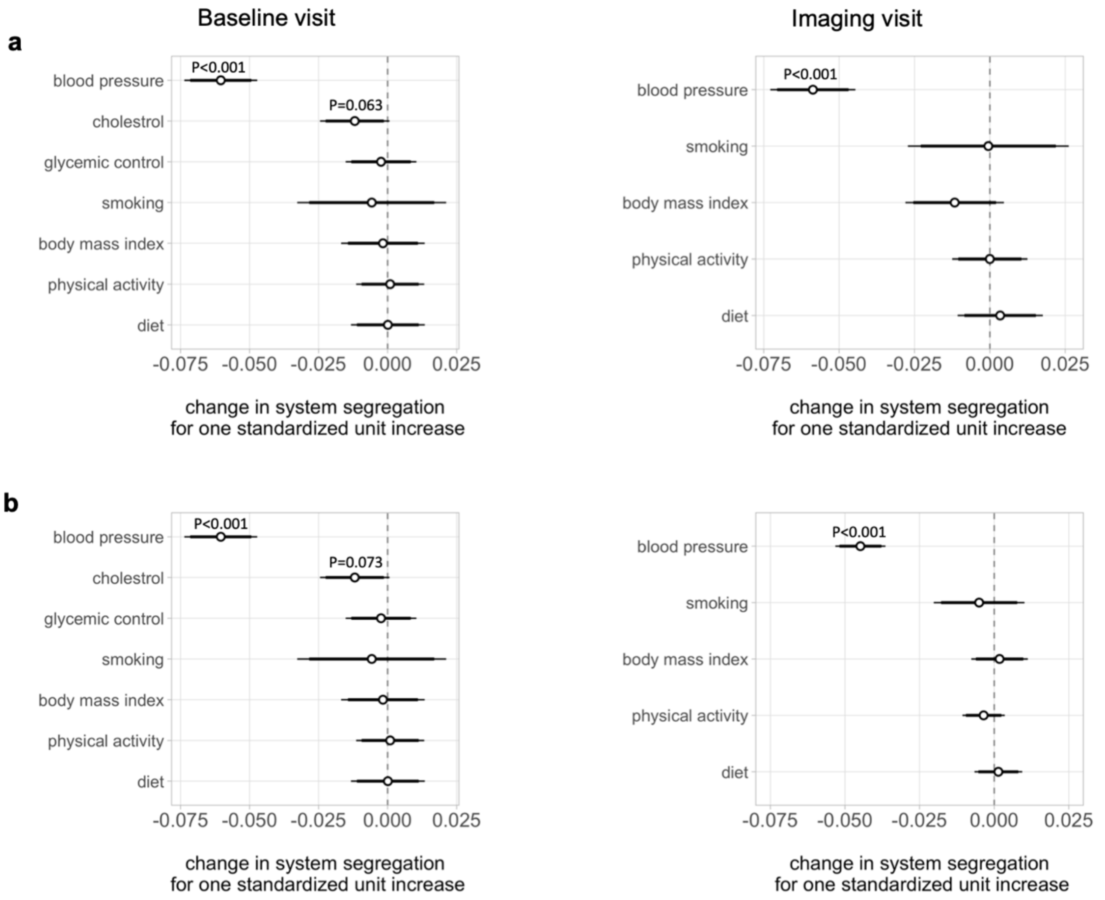


**Supplementary Fig. 4 Association between Life's Simple 7 (LS7) cardiovascular risk factors and system segregation**

Whisker plots show the association between continuous (standardized) measures of LS7 sub-scores and system segregation in **a** the imputed dataset (N = 16,635) or **b** non-imputed datasets (largest N = 16,084 for BMI at baseline visit and smallest N = 12,929 for diet at imaging visit) of the UK Biobank sample. All LS7 sub-scores had been acquired at the baseline (upper panel) or imaging visit (lower panel), except that blood has been drawn only at baseline (cholesterol and glycemic control). Statistical results were derived from multiple linear regression analyses including age, age$^2$, sex, education, grey matter volume, white matter hyperintensities volume, head motion during fMRI (i.e. frame-wise displacement) and assessment center as covariates. P-values < 0.1 are indicated, while Bonferroni-corrected P-values < 0.007 (0.05/7) are considered significant. Standardized beta coefficients are displayed together with 95% confidence intervals.

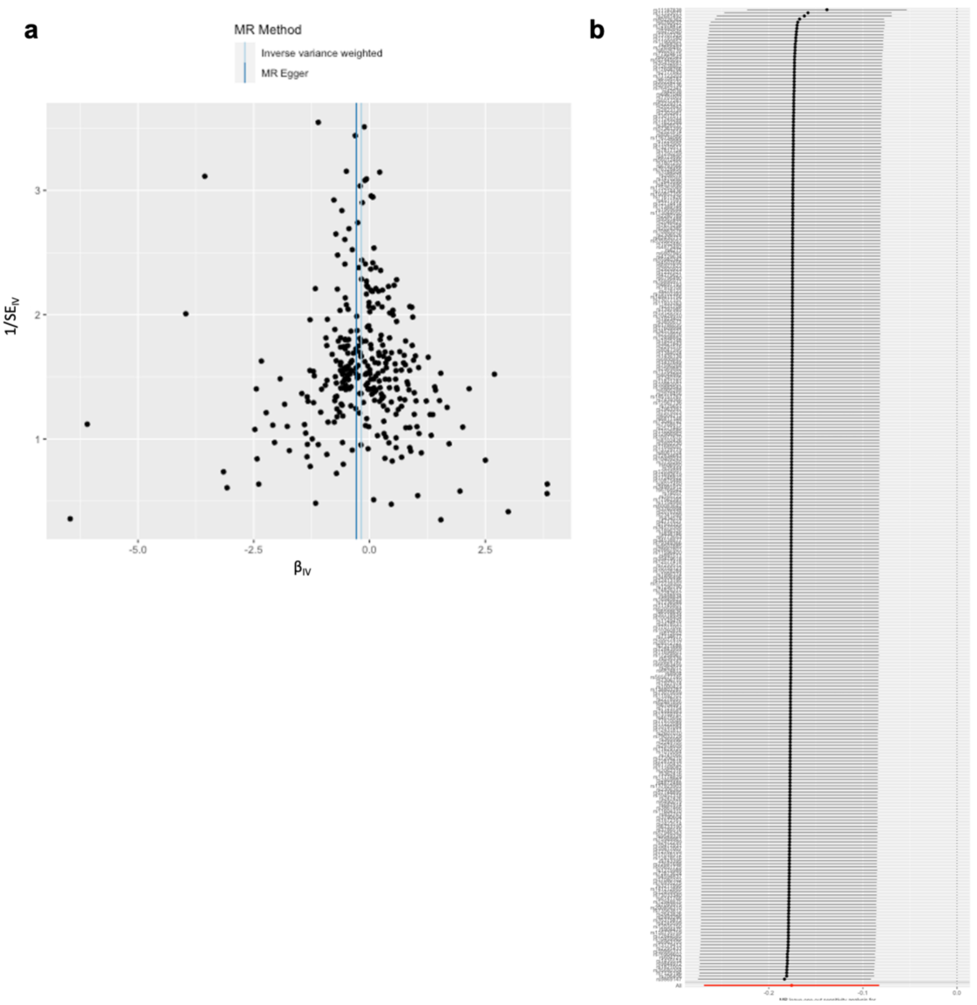


MR leave-one-out sensitivity analysis for systolic blood pressure on system segregation

**Supplementary Fig. 5 Quality assessment of Mendelian randomization results**

**a** The funnel plot shows the MR causal estimates (x-axis) against their precision (y-axis). Each data point corresponds to the effect of an individual genetic instrument of blood pressure on system segregation. The inverse-variance weighted (IVW) and MR-Egger causal effect estimates are also displayed. The funnel plot's symmetry indicates undirected pleiotropy meaning that weak instruments did not skew the results in one direction. **b** The leave-one-out analysis plot displays the IVW estimate after one instrument was removed at a time from the MR analysis. The last row (red color) indicates the causal estimate including all 312 instruments. MR effect sizes are shown together with 95% confidence intervals.

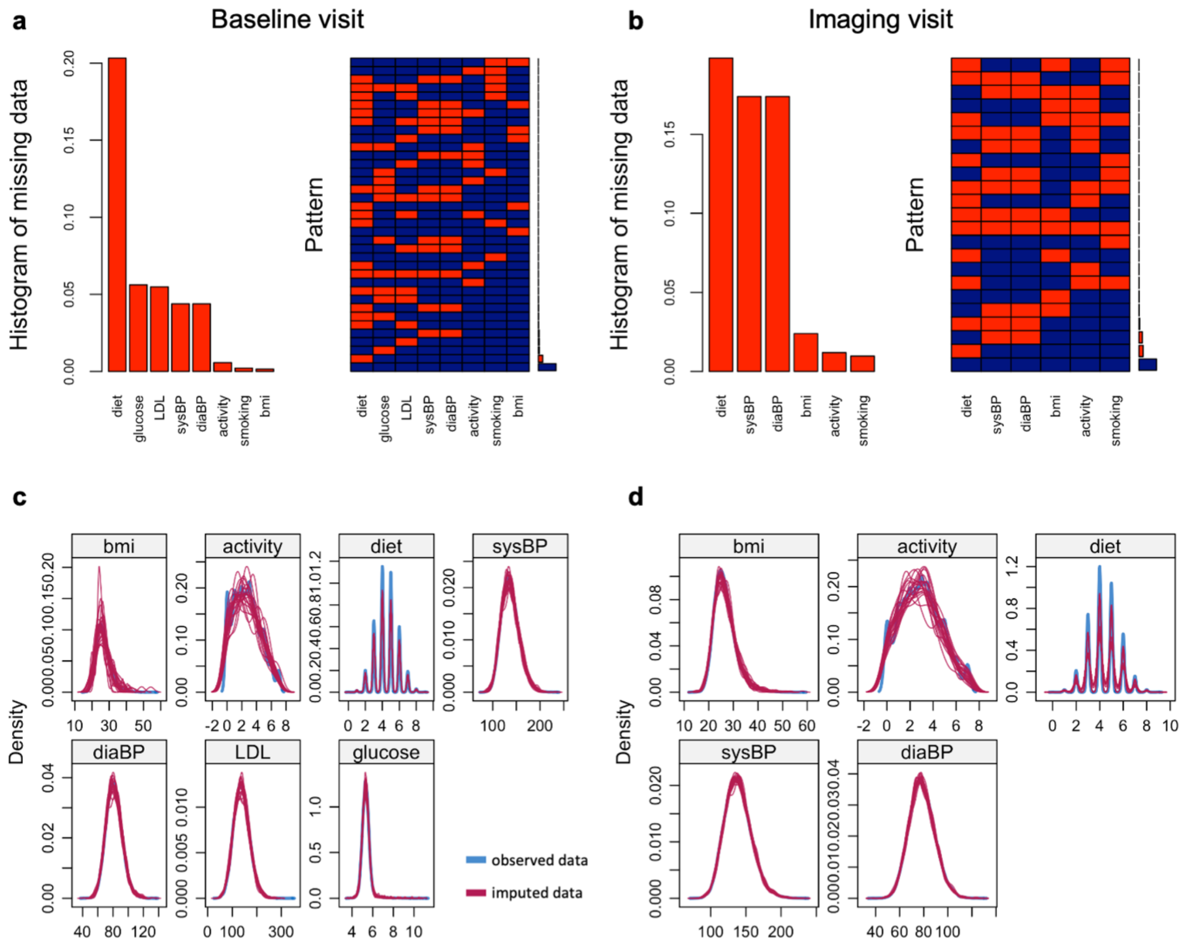


**Supplementary Fig. 6 Visualization of the data availability and quality of data imputation** Histograms show the percentage of missing data for each Life's Simple 7 (LS7) sub-score measured in **a** at the baseline and in **b** at the imaging visit of the UK Biobank. The diagrams to the right respectively illustrate the pattern of missing (red color) and observed data (blue color). Density plots display the distribution of the imputed data (red color) relative to the distribution of the observed data (blue color). Imputation has been performed by chained equations with 20 imputations and all remaining variables as predictors (R mice package).
